# Covid-19 vaccine safety in pregnancy, a nested case-control study in births from April 2021 to March 2022, England

**DOI:** 10.1101/2023.10.09.23296737

**Authors:** Anna A Mensah, Julia Stowe, Jennifer E Jardine, Freja C M Kirsebom, Tom Clare, Meaghan Kall, Helen Campbell, Jamie Lopez-Bernal, Nick Andrews

## Abstract

**Introduction:** Vaccine safety in pregnancy is always of paramount importance. Current evidence of COVID-19 vaccine safety in pregnancy has been reassuring with no association found with negative maternal and neonatal outcomes. However, very few safety studies are conducted on a national level and investigate dosage, timing of vaccination as well as vaccine manufacturer. To fill this knowledge gap, we conducted a population based COVID-19 vaccine safety evaluation in England, including timing of vaccination by trimester, dosage and vaccine manufacturer received in pregnancy.

**Method:** A matched case control study nested in a retrospective cohort where adverse maternal and neonatal pregnancy outcomes were compared across several COVID-19 vaccine exposures using conditional multivariable logistic regression, adjusting for a range of demographic and health characteristics. Eligible participants were identified from the national maternity services dataset (MSDS) and records were linked to hospital admission, national COVID-19 vaccine and COVID-19 testing databases. Matching criteria differed by outcome but included participant’s age and estimated week of conception.

**Results:** 514,013 pregnant individuals aged between 18 and 50 years were identified during the study period (births from 16th of April 2021-31st March 2022). Receiving at least one dose of COVID-19 vaccine during pregnancy conferred lower odds of giving birth to a baby who was low birthweight (aOR=0.86, 95% CI: 0.79 – 0.93), preterm (aOR=0.89, 95% CI: 0.85 - 0.92) or who had an Apgar score less than 7 at five mins of age (aOR=0.89, 95% CI: 0.80 - 0.98). There was no association between vaccination in pregnancy and stillbirth (aOR=0.90, 95% CI: 0.76 - 1.07), neonatal death (aOR=1.27, 95% CI: 0.91 - 1.77) perinatal death (aOR=0.98, 95% CI: 0.83 - 1.16), and maternal venous thromboembolism in pregnancy (aOR=0.82, 95% CI: 0.43 - 1.56). The odds of maternal admission to intensive care unit were lower in vaccinated pregnant women (aOR=0.85, 95% CI: 0.76 - 0.95).

**Conclusion:** COVID-19 vaccines are safe to use in pregnancy and they confer protection against SARS-CoV-2 infection which can lead to adverse outcomes for both the mother and the infant. Our findings generated important information to communicate to pregnant women and health professionals to support COVID-19 maternal vaccination programmes.

**What is already known on this topic:** Current evidence shows that COVID-19 vaccines are safe to use in pregnancy. However, few studies investigate the timing of vaccination in pregnancy including the first trimester for late pregnancy outcomes. Most studies are geographically limited, and few are population based allowing inclusion of participants representative of the country’s inhabitants.

**What this study adds:** This is the first population-based study in England investigating COVID-19 vaccine safety in pregnancy. We used the national maternity services dataset and national English health services data enabling inclusion of a huge numbers of participants across the country. As such, we were able to investigate relevant safety research questions such as the timing of vaccine administration in pregnancy by trimester and before pregnancy, the number of doses received and vaccine manufacturer.

**How this study might affect research, practice or policy:** This national study adds to further existing evidence demonstrating that all COVID-19 vaccines are safe to use in pregnancy at any point in time and gives pregnant individuals confidence in the COVID-19 maternal vaccine programme. We demonstrated that receiving multiple doses of COVID-19 vaccine in pregnancy is not associated with adverse pregnancy outcomes and additionally it was reassuring that there was no evidence of an increased risk by vaccine type.

## Introduction

Pregnant individuals are at increased risk of severe disease from SARS-CoV-2 infection compared to non-pregnant individuals putting both mothers and infants at risk of adverse outcomes during pregnancy and the postnatal period(1). Current knowledge indicates a higher risk of morbidity, intensive care unit (ICU) admission, ventilatory support, pre-eclampsia and preterm delivery in pregnant individuals with COVID-19 compared to uninfected pregnant individuals (2,3). During the early period of the pandemic the maternal mortality rate in unvaccinated pregnant individuals with COVID-19 was reported to be 0.8% with a ICU admission rate of 11.1% (4). Adverse outcomes in infants of SARS-CoV-2 infected mothers have also been documented with an increased risk of neonatal ICU admissions, stillbirth and neonatal mortality (5).

In the UK, starting 16 April 2021, pregnant individuals were encouraged to get COVID-19 vaccines when eligible based on their age and clinical risk group (6). On 2 December 2021, pregnant individuals were included in the UK’s priority COVID-19 vaccine distribution list (7). Oxford/AstraZeneca (ChAdOx1-S), Pfizer BioNTech (BNT162b2) or Moderna (mRNA-1273) are the most commonly used vaccines licensed in the UK (8).

COVID-19 vaccines have been shown to provide high levels of protection against severe COVID-19 in pregnancy (9,10). Vaccination against COVID-19 is therefore the most effective way to reduce adverse maternal and neonatal adverse outcomes due to SARS-CoV-2 infection in pregnancy.

Many studies and meta-analyses have found no increased risk of a range of adverse maternal, pregnancy or neonatal outcomes associated with COVID-19 vaccine administration in pregnancy including miscarriages, stillbirth or low birth weight and evidence of some more favourable outcomes such as in Apgar score or reduction in preterm birth and reduced risk of hospitalisation for COVID-19 related health issues in mothers and infants (11–15).

Our aim with this study is to further the knowledge of COVID-19 vaccine in pregnancy by investigating adverse pregnancy related, maternal and neonatal outcomes in pregnant individuals undertaking the first population-based approach using data from the English National Health Service (NHS). We aim to investigate vaccine safety stratified by the timing at which vaccine was received by gestational trimester, number of doses and type of vaccines administered.

## Methods

We conducted a matched case control study nested in a retrospective cohort covering all births from 16 April 2021 to 31 March 2022 to investigate adverse maternal and neonatal outcomes, comparing participants with the outcome (cases) and those without (controls) according to exposures to COVID-19 vaccine in pregnancy.

### Study population

The eligible population included all singleton pregnancies from individuals aged between 18 to 50 years at the time of pregnancy start with a delivery date between 16 April 2021 and 31 March 2022. Only records with a valid NHS number were included to allow linkage between datasets. (1)

The 16 April 2021 marks the beginning of the inclusion of pregnant individuals in the routine immunisation programme (health care workers or women with severe comorbidity would have been eligible earlier in the year), and we assume their eligibility for immunisation until the last day of the pregnancy. The end date is when free community COVID-19 testing ended so that detection of COVID-19 infections during pregnancy remains equally possible for all participants throughout the period.

In England, stillbirths are notifiable by law if occurring from 24 gestational weeks onwards. As such, the recording of early pregnancy outcomes (before 24 weeks) is not expected to be consistent across health care providers, hindering the ability to differentiate terminations from miscarriages and the confidence that such events would be recorded in a systematic manner. Because of the uncertainty on the type and scope of biases from early pregnancy outcomes, we have included only pregnancies of at least 24 gestational weeks in the analyses.

### Data sources and linkages

Participant were first identified from the Maternity Services Data Set (MSDS) antenatal booking records. MSDS collects records of each stage of the maternity service care pathway in NHS-funded maternity services across England. It includes information collected from the first antenatal appointment, diagnosis throughout pregnancy and outcome of deliveries for mothers and neonates.

Records from participants were linked to the Hospital Episode Statistics database (HES) which contains records of all admissions, outpatient appointments and emergency care attendances for patients admitted to NHS hospitals in England. To determine participants’ COVID-19 vaccine history we used the National Immunisation Management System (NIMS) database which collects vaccination data on national vaccine programmes including COVID-19 vaccination for the population registered with the NHS in England along with demographic information from all individuals residing in England. SARS-CoV-2 infections were identified through the national laboratory reporting system which captures routine laboratory data on infectious diseases and antimicrobial resistance. We extracted all positive COVID-19 test results (both PCR and LFT) from NHS laboratories and community testing.

All linkages were made using the unique NHS numbers from mothers. Deaths were identified using Office for National Statistics (ONS) data on all-cause mortality using infants NHS number for linkage. Where a valid NHS number was missing for the infant, they were excluded from the neonatal and perinatal deaths analyses as linkage to ONS deaths registration wasn’t possible. All data were extracted in November 2022. In the UK, all COVID-19 vaccines were given via NHS programme and, in England, over 99% of births occur within the NHS (16).

### Outcomes and case definition

Cases are defined as pregnant individuals experiencing any outcome of interest in pregnancy or infants born with any outcomes of interest. Stillbirth, low birthweight (below 2.5 kg), premature birth (before 37 weeks gestation) and venous thromboembolism (VTE) diagnosis outcomes used combination of the information found in both MSDS and HES datasets to enable comprehensive ascertainment of events (17). Maternal ICU admission was extracted solely from HES. Neonatal deaths (death of a live born baby before 28 days of age) were identified using Office for National Statistics (ONS) data on all-cause mortality. The baby’s Apgar score at five mins of age was extracted from MSDS.

Gestational length was calculated using firstly gestational age at birth, secondly last menstrual period and delivery date. Trimesters were defined as: trimester 1 up to day 83, trimester 2 starting at day 84 until day 188 and trimester 3 starting at day 189 up until 308 (maximum recorded gestational age in HES is 44 weeks).

### Vaccine exposures

The vaccinated cohort comprised of multiple levels of exposure as detailed in SUPPLEMENT TABLE 1. It included women who received a vaccine less than 3 months before their last menstrual period (LMP) or estimated start date of pregnancy, and those who received the vaccine in pregnancy. Vaccine exposures in pregnancy were further categorised by trimester of vaccination, dosage, and vaccine manufacturer (BNT162b2; mRNA-1273 and ChAdox1-S). The unexposed group comprised of unvaccinated participants and individuals who went on to receive the vaccine after delivery.

### Covariates

The data collected from MSDS consisted of information on current and past pregnancies and social circumstances (18). We extracted the following: complex social factors, disability, parity, gestational age at first appointment (categorised as 12 weeks, still within the first trimester, or during the second or third trimester).

Comorbidities were identified from SNOMED and ICD-10 codes included in the MSDS maternal records (**Supplement tables 6 & 7**). They were identified by a clinician and separated into categories: endocrine disease, hypertensive disease, cardiac disease, pregnancy related, other medical conditions, infection, coagulation disorder or other. An aggregate variable was developed to include conditions known to aggravate risks for the study outcomes. Immediate presentations (for example “fever”) were not included in the classification which was restricted to diagnoses. We combined mothers’ records of previous stillbirths and previous losses before 24 weeks of gestation to account for previous adverse outcomes.

We identified participants with a history of COVID-19 in pregnancy as SARS-CoV-2 infection was considered to be an important confounder in our study. Where participants had more than one positive test result, new episodes were defined by requiring an interval of at least 90 days. In the analysis COVID-19 infection in pregnancy was classified as positive if occurring in pregnancy and before the index date.

Using NIMS demographics’ information allowed us to complete data on ethnicity, NHS region, index of multiple deprivation quintiles (IMD), and vaccine delivery priority groups: healthcare worker, at risk of COVID-19 complications (which includes clinically extremely vulnerable (CEV), severely immunosuppressed and “at risk” status) on all patients including those not vaccinated (19,20).

### Statistical Methods

For infants controls were matched to cases by maternal age, gestational length in weeks (except for prematurity outcome), and estimated calendar week of conception (full details in **SUPPLEMENT TABLE 1**). Age matching was because maternal age has been associated with increased risk of preterm delivery, low birth weight and perinatal death. Gestation length matching was due to the strong association between this and infant outcomes. Week of pregnancy start was to account for any period confounding, for example this study took place during the COVID-19 pandemic period throughout which dramatic changes occurred in terms of behaviours (self-imposed or government imposed), healthcare access (fluctuation of healthcare services availability during epidemic and in between waves). The index date used to determine vaccination status was date of birth for the infant outcomes except for prematurity where the index date for controls was the gestational age of the case. This was done to minimize bias by allowing the same pregnancy duration in controls as cases for vaccination to be given. For this outcome, to further reduce bias, it was also necessary to separate out vaccinations within 14 days of the index date because it was clear that fewer vaccinations were given in the 14 days prior to birth in the premature cases giving an apparent protective effect of vaccination.

Because of the strong correlation between low birthweight and gestational age, only pregnancies reaching 37 weeks were include in those analyses.

For maternal outcomes matching was the same as for infants, but the index date used to determine vaccination status was the date of the event in the case.

Up to 5 controls were matched to each case without replacement. This was done separately for each outcome (**SUPPLEMENT TABLE 2**). **SUPPLEMENT TABLE 1** illustrates in detail the matching process undertaken in the analyses.

We describe covariates for matched cases and controls by outcome status and assessed differences in covariate distribution between cases and controls using Chi-square or Kruskal–Wallis tests.

We used conditional multivariable logistic regression to assess the odds of each outcome by vaccination exposure. Models were adjusted for the following predefined covariates: age as a continuous variable (if it was not a matching criteria), ethnicity, IMD-5, comorbidities, complex social factors, disability, NHS worker, at risk of complication from COVID-19 disease, previous adverse pregnancy outcomes, COVID-19 infection during pregnancy. Odds ratios with 95% confidence intervals are reported for each model.

We used Microsoft SQL Server Management Studio 18 for data curation and linkages across datasets and STATA 17 for statistical analyses.

## Results

After data linkage and cleaning, we identified 514,013 pregnant women from 126 Hospital Trusts across England who gave birth during the study period of which 12,213 were not included in the analyses as the pregnancy ended before reaching 24 weeks (**SUPPLEMENT FIGURE 1**).

The median age of participants, calculated at their first antenatal booking appointment, was 30 years old (Inter Quartile Range (IQR) 25%-75%: 27 – 34 years). Prior to matching, there were significant differences between those vaccinated and not vaccinated for all covariates except region (TABLE 1). Vaccinated women were more likely to be older, of white ethnicity, and live in areas with lower levels of socioeconomic deprivation. There were also more likely to be in other groups targeted for vaccination including those with comorbidities, at risk of COVID-19 complications and health and care workers.

**Table 1:** Demographic data on pregnant participants by exposure status and P-Value.

| Risk factors / exposure |  | Vaccinated during pregnancy |  | P-value |
| --- | --- | --- | --- | --- |
|  |  | No | Yes |  |
| **Age groups in Years** |  | **330,347** | **182,477** |  |
|  | *18 to 24* | 63,557 (19.24) | 16,545 (9.07) | < 0.001** |
|  | *25 to 29* | 101,846 (30.83) | 42,201 (23.13) |  |
|  | *30 to 34* | 104,243 (31.56) | 72,343 (39.64) |  |
|  | *35 to 39* | 49,935 (15.12) | 42,277 (23.17) |  |
|  | *40 to 44* | 10,091 (3.05) | 8,704 (4.77) |  |
|  | *45 to 50* | 675 (0.20) | 407 (0.22) |  |
| **Ethnicity** |  | **325,590** | **178,867** |  |
|  | *White* | 239,668 (73.61) | 143,661 (80.32) | < 0.001** |
|  | *Mixed Multiple Ethnic groups* | 5,713 (1.75) | 2,150 (1.20) |  |
|  | *Black/Black British* | 21,193 (6.51) | 4,986 (2.79) |  |
|  | *Asian/Asian British* | 46,527 (14.29) | 23,263 (13.01) |  |
|  | *Any other ethnic groups* | 12,489 (3.84) | 4,807 (2.69) |  |
| **Region** |  | **323,604** | **179,526** |  |
|  | *East Midlands* | 26,561 (8.21) | 14,842 (8.27) | < 0.166** |
|  | *East of England* | 38,690 (11.96) | 21,602 (12.03) |  |
|  | *London* | 58,375 (18.04) | 30,032 (16.73) |  |
|  | *North East* | 13,750 (4.25) | 7,556 (4.21) |  |
|  | *North West* | 44,062 (13.62) | 21,596 (12.03) |  |
|  | *South East* | 46,543 (14.38) | 31,970 (17.81) |  |
|  | *South West* | 25,718 (7.95) | 19,084 (10.63) |  |
|  | *West Midlands* | 37,670 (11.64) | 16,498 (9.19) |  |
|  | *Yorkshire and Humber* | 32,235 (9.96) | 16,346 (9.11) |  |
| **Index of Multiple Deprivation** |  | **323,604** | **179,526** |  |
|  | *1 Most Deprived* | 92,575 (28.61) | 29,995 (16.71) | < 0.001** |
|  | *2* | 74,153 (22.91) | 33,982 (18.93) |  |
|  | *3* | 60,906 (18.82) | 37,022 (20.62) |  |
|  | *4* | 52,428 (16.20) | 38,601 (21.50) |  |
|  | *5 Least Deprived* | 43,542 (13.46) | 39,926 (22.24) |  |
| **Complex social factors** |  | **309,785** | **175,188** |  |
|  | *No* | 260,614 (84.13) | 159,786 (91.21) | < 0.001* |
|  | *Yes* | 49,171 (15.87) | 15,402 (8.79) |  |
| **Disability** |  | **273,807** | **150,936** |  |
|  | *No* | 264,694 (96.67) | 144,744 (95.90) | < 0.001* |
|  | *Yes* | 9,113 (3.33) | 6,192 (4.10) |  |
| **Parity** |  | **331,431** | **182,582** |  |
|  | *Nulliparous* | 128,026 (38.63) | 77,422 (42.40) | < 0.001* |
|  | *Multiparous* | 203,405 (61.37) | 105,160 (57.60) |  |
| **Gestational age at first appointment** |  | **329,700** | **182,211** |  |
|  | *< 12 weeks* | 284,018 (86.14) | 166,328 (91.28) |  |
|  | *1st Trimester* | 16,899 (5.13) | 7,336 (4.03) | < 0.001** |
|  | *2nd Trimester* | 22,699 (6.88) | 6,993 (3.84) |  |
|  | *3rd Trimester* | 6,084 (1.85) | 1,554 (0.85) |  |
| **Previous adverse outcome** |  | **331,431** | **182,582** |  |
|  | *No* | 202,883 (61.21) | 119,454 (65.42) | < 0.001* |
|  | *Yes* | 128,548 (38.79) | 63,128 (34.58) |  |
| **Comorbidities** |  | 331431 | 182582 |  |
|  | *No* | 315,747 (95.27) | 172,768 (94.62) | < 0.001* |
|  | *Yes* | 15,684 (4.73) | 9,814 (5.38) |  |
| **NHS worker** |  | **330,010** | **182,485** |  |
|  | *No* | 309,453 (93.77) | 162,737 (89.18) | < 0.001* |
|  | *Yes* | 20,557 (6.23) | 19,748 (10.82) |  |
| **At risk of COVID-19 complications** |  | **330,010** | **182,485** |  |
|  | *No* | 271,186 (82.18) | 142,511 (78.09) |  |
|  | *Yes* | 58,824 (17.82) | 39,974 (21.91) | < 0.001* |
| **COVID-19 Infection in Pregnancy** |  | **331,431** | **182,582** |  |
|  | *No positive test in Pregnancy* | 289,359 (87.31) | 154,789 (84.78) | < 0.001* |
|  | *Positive test during pregnancy* | 42,072 (12.69) | 27,793 (15.22) |  |

\*P-Value from Chi2-test
\*\* P-value from Kruskal–Wallis test (Chi2 with ties)

4.4% (22,758) of infants with a birth weight below 2.5kg during the study period. 40,122 (7.8%) babies were born before 37 weeks gestation during the study period. During the study period, we identified 1,909 (0.37%) stillbirths for babies born after a minimum of 24 gestational weeks. They were 539 (1.3‰) live born babies who died within 28 days of birth during the study period. Most babies had a normal Apgar score at five mins after birth with 8.9‰ (4,562) of babies scoring below7. Less than 1% (4,895) of participants were admitted to ICU, while 295 (0.06%) had VTE during their pregnancy or 42 days within giving birth during the study period.

The peak of vaccination in pregnancy was in May and June 2021 when most pregnant individuals were prioritised for vaccination while coincidently reaching the eligible age for routine immunisation (the below 40 years age group). (**FIGURE 1**)

**Figure 1:**
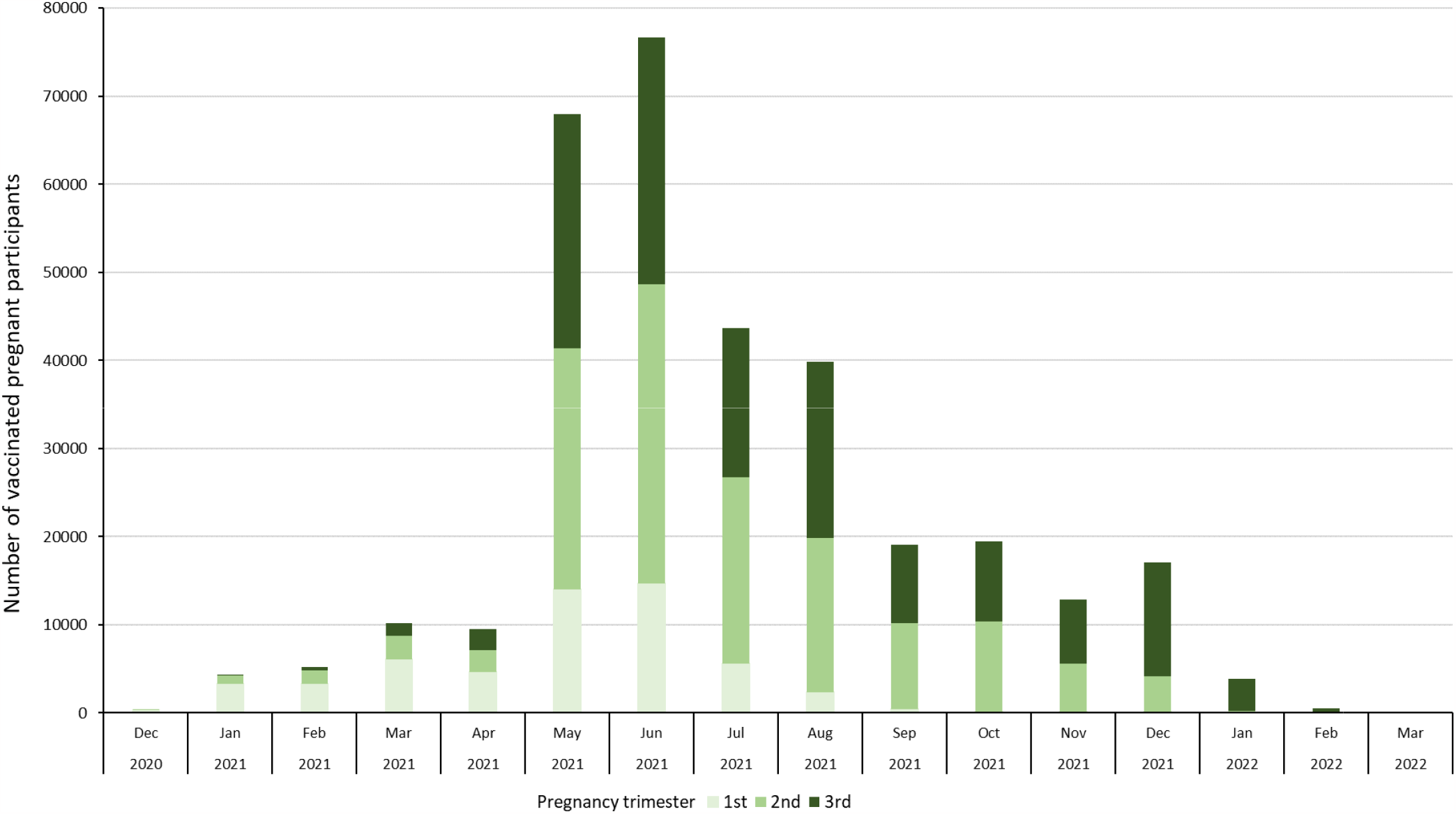
Number of vaccinated participants during the study period per trimester when first vaccine dose in pregnancy was received

### Matching

The number of controls matched to each case is given in **SUPPLEMENT TABLE 2**. For most outcomes a large majority had the full number of 5 controls matched.

### Low birthweight, preterm births and low Apgar score

Overall, births of babies with low birthweight were less likely to occur in women who received at least one dose of COVID-19 vaccine in pregnancy (adjusted Odds Ratio (aOR)=0.86, 95% Confidence Interval (CI): 0.79 – 0.93) or two doses (aOR=0.84, 95% CI: 0.76 – 0.93) in pregnancy (**TABLE 2**). In subgroup analyses, similar results were seen regardless of trimester and vaccine used, although results for first trimester or immediately pre-pregnancy or if vaccinated with ChAdOx1 did not reach statistical significance due to low numbers (**TABLE 2**).

**Table 2:**
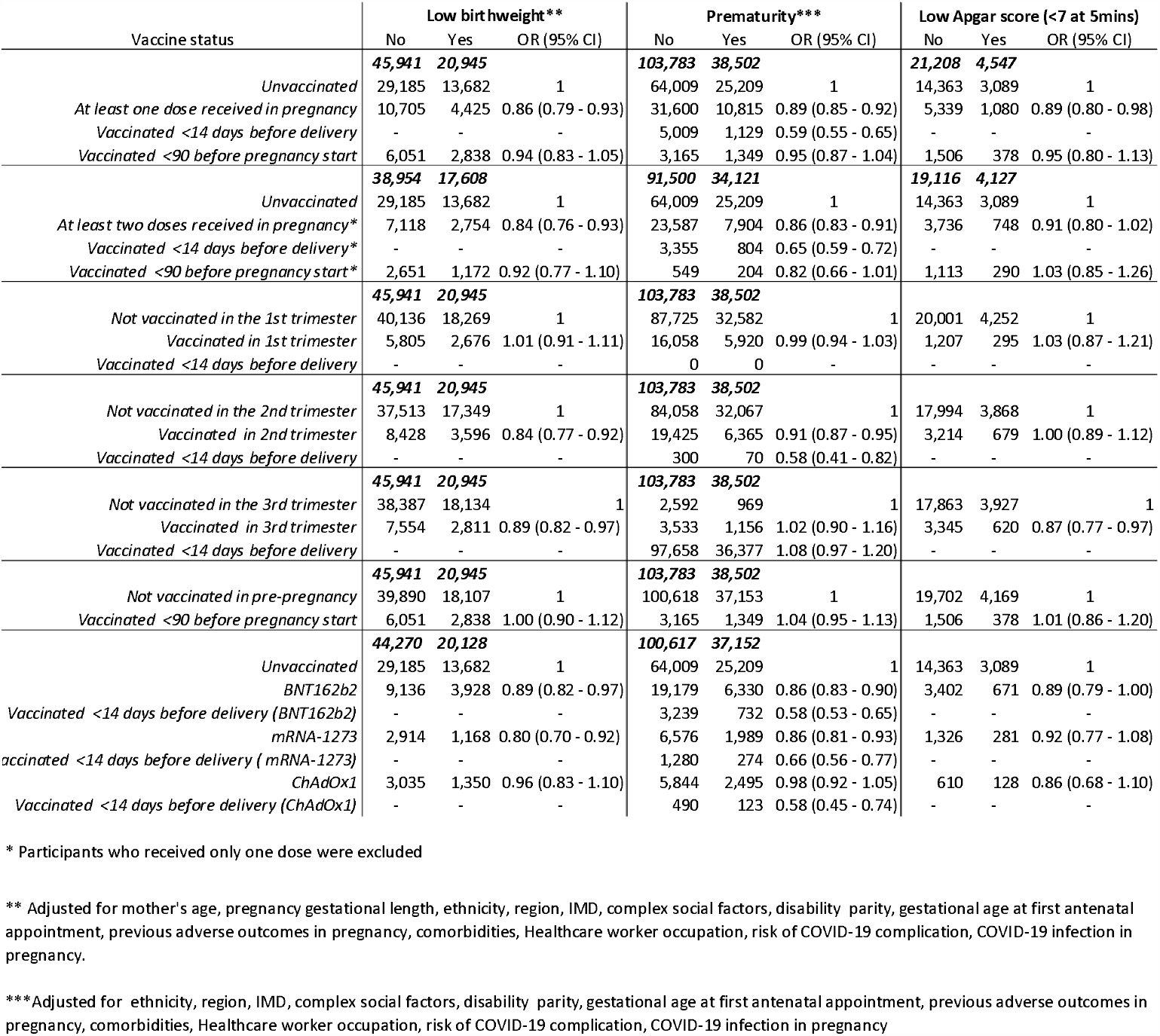
Adjusted odd ratios from conditional logistic regression.

Odds of preterm birth were lower in individuals vaccinated with at least one dose (aOR=0.85, 95% CI: 0.85 - 0.92), in their second trimester (aOR=0.91, 95% CI: 0.87 - 0.95) and with BNT162b2 or mRNA-1273: (aOR=0.86, 95% CI: 0.83 - 0.90) and aOR=0.86, 95% CI: 0.81 - 0.93) respectively. Individuals vaccinated within 14 days of the date of birth(case) or index date(control) had even lower odds (aOR=0.59, 95% CI: 0.55 - 0.65) (**TABLE 2**).

A small protective effect against Apgar score lower than 7 at five mins after birth was observed in babies whose mother’s received at least one dose of vaccine in pregnancy (aOR=0.89, 95% CI: 0.80 - 0.98) and in those vaccinated with BNT162b2 (aOR=0.89, 95% CI: 0.79 - 1.00).

### Stillbirth, neonatal and perinatal deaths

We found no association between stillbirth and individuals receiving COVID-19 vaccine in pregnancy as all CI include 1 (**TABLE 3**). There was marginal protection offered from vaccine received within 90 days prior to the start of the pregnancy (aOR=0.79, 95% CI: 0.63 – 1.00).

**Table 3:**
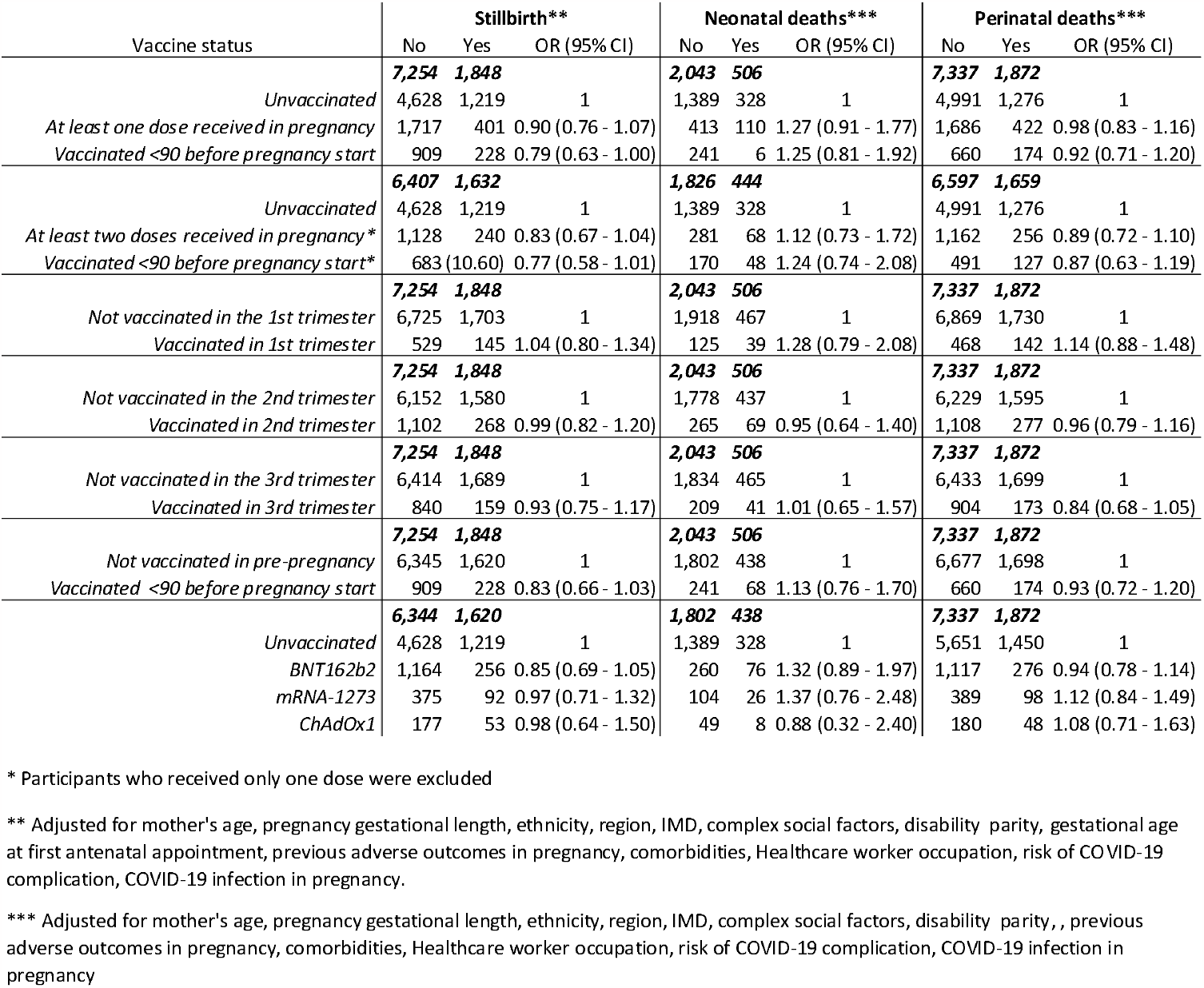
Adjusted odd ratios from conditional logistic regression.

As previously indicated in the methods, infants with no NHS number were excluded from the neonatal and perinatal analyses (**SUPPLEMENT FIGURE 1**). We found no association between deaths of an infant within 28 days of birth and mother’s vaccine exposure in pregnancy (**TABLE 3**), with similar results for perinatal deaths as all CI include 1.

### Venous thromboembolism (VTE) and ICU admission in pregnancy and during the perinatal period

Pregnant individuals who received at least one dose or at least two doses of COVID-19 vaccine within 90 days of the pregnancy start had lower odds of developing VTE in pregnancy (aOR=0.55, 95% CI: 0.32 - 0.94) and (aOR=0.45, 95% CI: 0.20 - 0.98) respectively (**TABLE 4**). This was also observed in those who received at least one dose of BNT162b2 (aOR=0.33, 95% CI: 0.11 - 0.99). The results were not statistically significant for other exposures.

**Table 4:**
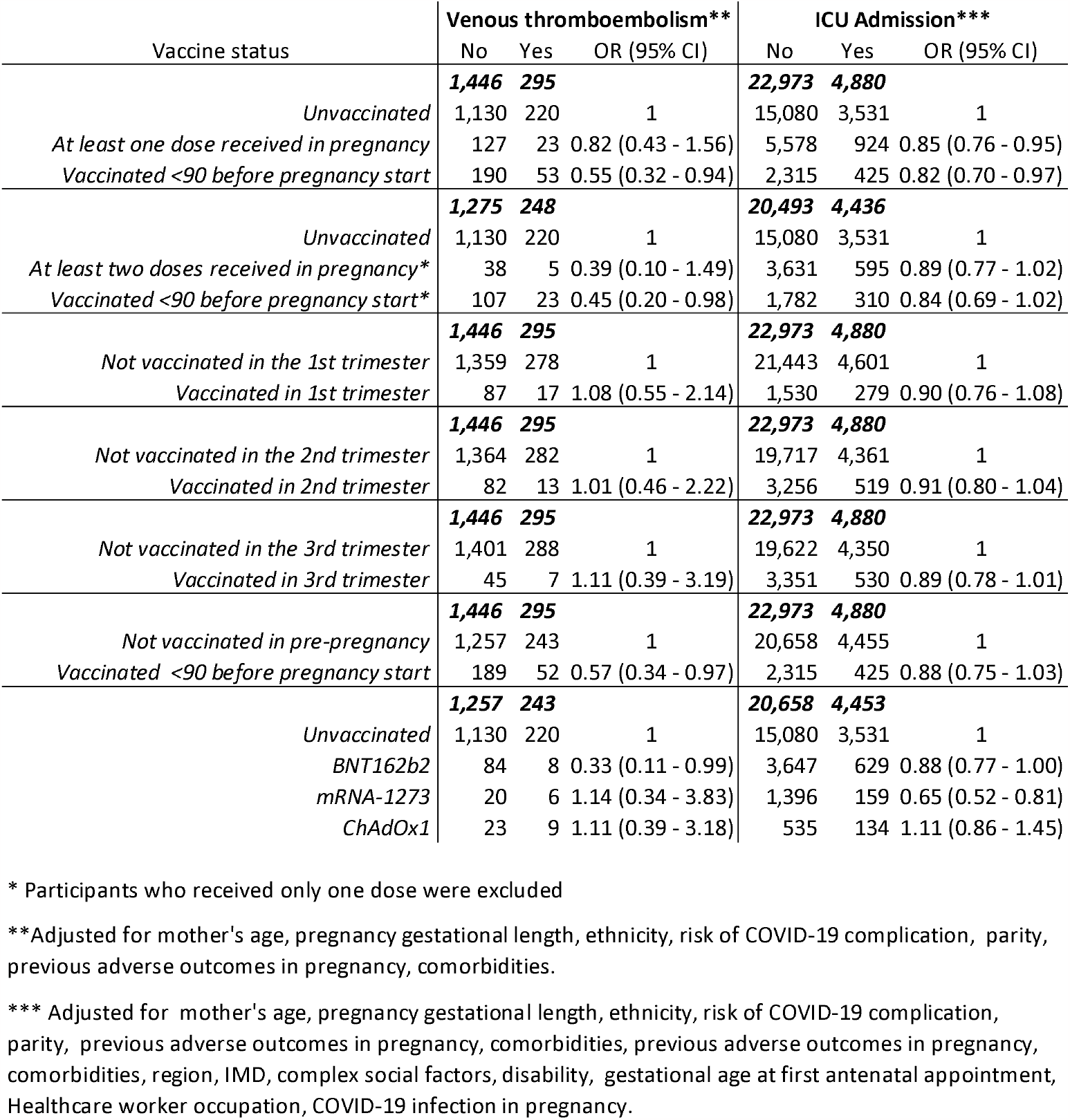
Adjusted odd ratios from conditional logistic regression.

Finally, only those who received at least one dose of vaccine during pregnancy, within 90 days of pregnancy start or received at least one dose of mRNA-1273 had lower odds of ICU admission (aOR=0.85, 95% CI: 0.76 - 0.96), (aOR=0.82, 95% CI: 0.70 - 0.97) and (aOR=0.65, 95% CI: 0.52 - 0.81) respectively (**TABLE 4**).

## Discussion

Overall, we found that COVID-19 vaccines are safe to use in pregnancy with no evidence that exposed groups are at any increased risk of maternal, neonatal or pregnancy adverse outcomes. We found that receiving at least one dose of vaccine in pregnancy gave possible protective effects against low birthweight, preterm birth, Apgar results below 7 five minutes after birth and ICU admission in pregnancy or during the postpartum period. We found no association between stillbirth, neonatal deaths, perinatal deaths and VTE vaccine exposure in pregnancy. Finally, results stratified by timing of vaccination indicated no evidence of a safety risk for COVID-19 vaccines administered in pregnancy during the first, second or third trimester.

Our results are consistent with other studies from England where a cohort study including individuals who gave birth at a single hospital in London between March 2020 and July 2021 found no differences in stillbirth, foetal abnormalities, postpartum haemorrhage and infants born small for gestational age, when comparing unvaccinated women with those that received at least one dose of vaccine during pregnancy during their second and/or third pregnancy trimester (P>.05 for all)(21). A small protective effects against some outcomes in the exposed groups might be the result of a protection against SARS-CoV-2 infection which is known to increase the risk of these outcomes; while some of this effect might also be attributed to the healthy vaccinee effect (22). This bias might manifest through women who are unwell in pregnancy missing the opportunity to get vaccinated or due to generally poorer health in those not choosing to get vaccinated based on factors we did not measure and adjust for in the analysis (residual confounding).

### This protective effect could also be the result of protection against infection that conferred COVID-19 vaccine

Vaccination close to the end of pregnancy was also less likely as demonstrated in the analysis of prematurity with an apparent protective effect in this period. COVID-19 vaccine safety in pregnancy has also been assessed in various other countries and similarly to our findings, results have either showed no difference between vaccinated and unvaccinated groups or some protective effect against adverse outcomes in pregnancy in the vaccinated (15,23–25). A study in Switzerland which looked at time of vaccine administration and occurrence of adverse pregnancy outcomes such as early pregnancy loss and preterm deliveries found similar rates in COVID-19 vaccinated individuals when compared to historical data on background risks in the obstetric population (26). Although not investigated in this study, other studies show evidence that reactogenicity from the administration of COVID-19 vaccines in pregnancy is no cause for concern and further strengthens the safety profile of COVID-19 vaccines (27).

In addition to being safe, vaccines in pregnancy have been shown to be highly effective against mild disease and hospitalisation in mothers whilst also offering protection from symptomatic disease and hospitalisation in infants of vaccinated women (9,10,28,29). Furthermore, if a booster dose is administered in the later stages of pregnancy it has the potential to increase maternal and neonatal immunity against SARS-CoV-2(30). By protecting expectant individuals against COVID-19, vaccination will also protect pregnant individuals against disease associated increased risks of preeclampsia, preterm birth and low birthweight delivery (31).

Our findings that unvaccinated participants tended to be of non-white ethnicity and of lower social economic background were also reported in a single site cohort study corroborating our finding at the national level (21). It is therefore important to effectively communicate the safety of COVID-19 vaccine in pregnancy to enhance coverage with the aim of reducing disease burden and health inequalities.

One of the strengths of this study is the linkage of a wide range of national datasets; maternity services, hospital admission records, national immunisation and covid-19 testing databases. We were able to include many participants from across all parts of England with an estimated 86% (514,013/597,708) coverage when compared to Office for National Statistics births from 2021(16).This study is one of the few that look at vaccine safety stratified by time and number of doses received during pregnancy while also considering the vaccine’s manufacturer.

A limitation of this study was missing infant NHS numbers which led to a substantive part of the cohort being removed for the neonatal and perinatal deaths analyses. However, the overall mortality ratio in our study sample was very similar to the reported ONS estimates for 2021 (32). NHS numbers were predominantly missing from the most recent pregnancy records illustrating the delay in this data being made available. Additionally, we could not include maternal deaths in our analyses due to data missing for a small but substantial number of pregnancies.

Throughout the pandemic, the succession of circulating SARS-CoV-2 variants have had an impact on COVID-19 disease severity. In fact, in a population where vaccine coverage and prior infection are high, disease severity in pregnancy from the Omicron variant was less than previously observed from infection with Delta variant (33). Further studies should focus on establishing the current burden of COVID-19 disease in pregnancy to help review current vaccination guidelines while we continue assessing the safety profile of novel vaccines tailored to tackle new arising strains. This process would need to be supported by a robust and timely data collection and reporting withing the biological limits of pregnancy timings.

## Conclusion

We found that COVID-19 vaccines are safe to use in pregnancy with no observed increase in adverse maternal and neonatal outcomes following administration in pregnancy. They confer protection against SARS-CoV-2 infection which can lead to adverse outcomes for both the mother and the infant. Our findings generated important information to communicate to pregnant women and health professionals to support COVID-19 maternal vaccine programmes. The current burden of COVID-19 disease in pregnancy should be assessed.

## Data Availability

This work is carried out under Regulation 3 of The Health Service (Control of Patient Information; Secretary of State for Health, 2002) using patient identification information without individual patient consent as part of the UKHSA legal requirement for public health surveillance and monitoring of vaccines. As such, authors cannot make the underlying dataset publicly available for ethical and legal reasons. However, all the data used for this analysis is included as aggregated data in the manuscript tables and appendix. Applications for relevant anonymised data should be submitted to the UKHSA Office for Data Release at https://www.gov.uk/government/publications/accessing-ukhsa-protected-data.

## Authors’ Contributions

JLB, NA, HC, JS, AAM conceptualised the study. JJ reviewed the SNOMED and ICD-10 codes. FCK provided the COVID-19 testing data. TC and MK provided the ONS all mortality cause data. AAM curated the data. NA designed the analysis plan and AAM conducted the formal analysis assisted by NA. JS accessed and verified the data. AAM, JS wrote the original draft of the manuscript. HC wrote the introduction. All co-authors reviewed the manuscript and were responsible for the decision to submit the manuscript.

## Ethics Committee Approval

Surveillance of COVID-19 testing and vaccination is undertaken under Regulation 3 of The Health Service (Control of Patient Information) Regulations 2002 to collect confidential patient information (www.legislation.gov.uk/uksi/2002/1438/regulation/3/made) under Sections 3(i) (a) to (c), 3(i)(d) (i) and (ii) and 3(3). The study protocol was subject to an internal review by the UK Health Security Agency Research Ethics and Governance Group and was found to be fully compliant with all regulatory requirements. As no regulatory issues were identified, and ethical review is not a requirement for this type of work, it was decided that a full ethical review would not be necessary.

## References

1. Allotey J, Fernandez S, Bonet M, Stallings E, Yap M, Kew T, et al. Clinical manifestations, risk factors, and maternal and perinatal outcomes of coronavirus disease 2019 in pregnancy: living systematic review and meta-analysis. BMJ. 2020 Sep 1;370:m3320.

2. Boettcher LB, Metz TD. Maternal and neonatal outcomes following SARS-CoV-2 infection. Semin Fetal Neonatal Med. 2023 Feb;28(1):101428.

3. Mascio DD, Sen C, Saccone G, Galindo A, Grünebaum A, Yoshimatsu J, et al. Risk factors associated with adverse fetal outcomes in pregnancies affected by Coronavirus disease 2019 (COVID-19): a secondary analysis of the WAPM study on COVID-19. J Perinat Med. 2020 Nov 1;48(9):950–8.

4. Covid-19 TW (World A of PMWG on. Maternal and perinatal outcomes of pregnant women with SARS-CoV-2 infection. Ultrasound Obstet Gynecol. 2021;57(2):232–41.

5. Papapanou M, Papaioannou M, Petta A, Routsi E, Farmaki M, Vlahos N, et al. Maternal and Neonatal Characteristics and Outcomes of COVID-19 in Pregnancy: An Overview of Systematic Reviews. Int J Environ Res Public Health. 2021 Jan 12;18(2):596.

6. GOV.UK [Internet]. [cited 2023 Jul 15]. JCVI issues new advice on COVID-19 vaccination for pregnant women. Available from: https://www.gov.uk/government/news/jcvi-issues-new-advice-on-covid-19-vaccination-for-pregnant-women

7. GOV.UK [Internet]. 2021 [cited 2023 Jul 15]. Pregnant women urged to come forward for COVID-19 vaccination. Available from: https://www.gov.uk/government/news/pregnant-women-urged-to-come-forward-for-covid-19-vaccination

8. GOV.UK [Internet]. 2023 [cited 2023 Jul 18]. COVID-19: the green book, chapter 14a. Available from: https://www.gov.uk/government/publications/covid-19-the-green-book-chapter-14a

9. Kirsebom FCM, Andrews N, Mensah AA, Stowe J, Ladhani SN, Ramsay M, et al. Vaccine effectiveness against mild and severe disease in pregnant mothers and their infants in England [Internet]. medRxiv; 2023 [cited 2023 Jul 14]. p. 2023.06.07.23290978. Available from: https://www.medrxiv.org/content/10.1101/2023.06.07.23290978v1

10. Villar J, Conti CPS, Gunier RB, Ariff S, Craik R, Cavoretto PI, et al. Pregnancy outcomes and vaccine effectiveness during the period of omicron as the variant of concern, INTERCOVID-2022: a multinational, observational study. The Lancet. 2023 Feb 11;401(10375):447–57.

11. Calvert C, Carruthers J, Denny C, Donaghy J, Hillman S, Hopcroft LEM, et al. A populationbased matched cohort study of early pregnancy outcomes following COVID-19 vaccination and SARS-CoV-2 infection. Nat Commun. 2022 Oct 17;13(1):6124.

12. Prabhu M, Riley LE. Coronavirus Disease 2019 (COVID-19) Vaccination in Pregnancy. Obstet Gynecol. 2023 Mar 2;141(3):473–82.

13. Rahmati M, Yon DK, Lee SW, Butler L, Koyanagi A, Jacob L, et al. Effects of COVID-19 vaccination during pregnancy on SARS-CoV-2 infection and maternal and neonatal outcomes: A systematic review and meta-analysis. Rev Med Virol. 2023 May;33(3):e2434.

14. Shafiee A, Kohandel Gargari O, Teymouri Athar MM, Fathi H, Ghaemi M, Mozhgani SH. COVID-19 vaccination during pregnancy: a systematic review and meta-analysis. BMC Pregnancy Childbirth. 2023 Jan 20;23:45.

15. Prasad S, Kalafat E, Blakeway H, Townsend R, O’Brien P, Morris E, et al. Systematic review and meta-analysis of the effectiveness and perinatal outcomes of COVID-19 vaccination in pregnancy. Nat Commun. 2022 May 10;13(1):2414.

16. Provisional births in England and Wales - Office for National Statistics [Internet]. [cited 2023 Sep 14]. Available from: https://www.ons.gov.uk/peoplepopulationandcommunity/birthsdeathsandmarriages/livebirths/articles/provisionalbirthsinenglandandwales/2021

17. NHS Digital [Internet]. [cited 2023 Jul 14]. Hospital Episode Statistics (HES). Available from: https://digital.nhs.uk/data-and-information/data-tools-and-services/data-services/hospital-episode-statistics

18. NHS Digital [Internet]. [cited 2023 Jul 14]. Implementing the Maternity Services Data Set (MSDS) v2.0 tools and guidance. Available from: https://digital.nhs.uk/data-and-information/data-collections-and-data-sets/data-sets/maternity-services-data-set/tools-and-guidance

19. NHS Englandlll» National Vaccination Programmes [Internet]. [cited 2023 Jul 17]. Available from: https://www.england.nhs.uk/contact-us/privacy-notice/national-flu-vaccination-programme/

20. NHS Digital [Internet]. [cited 2023 Jul 17]. Cohorting as a Service (CaaS). Available from: https://digital.nhs.uk/services/cohorting-as-a-service-caas

21. Blakeway H, Prasad S, Kalafat E, Heath PT, Ladhani SN, Le Doare K, et al. COVID-19 vaccination during pregnancy: coverage and safety. Am J Obstet Gynecol. 2022 Feb;226(2):236.e1-236.e14.

22. Shrank WH, Patrick AR, Alan Brookhart M. Healthy User and Related Biases in Observational Studies of Preventive Interventions: A Primer for Physicians. J Gen Intern Med. 2011 May;26(5):546–50.

23. Covas DT, de Jesus Lopes de Abreu A, Zampirolli Dias C, Vansan Ferreira R, Gonçalves Pereira R, Silva Julian G. Adverse events of COVID-19 vaccines in pregnant and postpartum women in Brazil: A cross-sectional study. PLOS ONE. 2023 Jan 13;18(1):e0280284.

24. Lipkind HS, Vazquez-Benitez G, DeSilva M, Vesco KK, Ackerman-Banks C, Zhu J, et al. Receipt of COVID-19 Vaccine During Pregnancy and Preterm or Small-for-Gestational-Age at Birth — Eight Integrated Health Care Organizations, United States, December 15, 2020–July 22, 2021. MMWR Morb Mortal Wkly Rep. 2022 Jan 7;71(1):26–30.

25. Shimabukuro TT, Kim SY, Myers TR, Moro PL, Oduyebo T, Panagiotakopoulos L, et al. Preliminary Findings of mRNA Covid-19 Vaccine Safety in Pregnant Persons. N Engl J Med. 2021 Jun 17;384(24):2273–82.

26. Favre G, Maisonneuve E, Pomar L, Winterfeld U, Daire C, Martinez de Tejada B, et al. COVID-19 mRNA vaccine in pregnancy: Results of the Swiss COVI-PREG registry, an observational prospective cohort study. Lancet Reg Health Eur. 2022 Jul;18:100410.

27. Sadarangani M, Soe P, Shulha HP, Valiquette L, Vanderkooi OG, Kellner JD, et al. Safety of COVID-19 vaccines in pregnancy: a Canadian National Vaccine Safety (CANVAS) network cohort study. Lancet Infect Dis. 2022 Nov;22(11):1553–64.

28. Maternal Vaccination and Risk of Hospitalization for Covid-19 among Infants | NEJM [Internet]. [cited 2023 Jul 14]. Available from: https://www.nejm.org/doi/full/10.1056/nejmoa2204399

29. Immunogenicity of COVID-19 mRNA Vaccines in Pregnant and Lactating Women | Breastfeeding | JAMA | JAMA Network [Internet]. [cited 2023 Jul 14]. Available from: https://jamanetwork.com/journals/jama/fullarticle/2780202

30. Atyeo C, Shook LL, Nziza N, Deriso EA, Muir C, Baez AM, et al. COVID-19 booster dose induces robust antibody response in pregnant, lactating, and nonpregnant women. Am J Obstet Gynecol. 2023 Jan;228(1):68.e1-68.e12.

31. Wei SQ, Bilodeau-Bertrand M, Liu S, Auger N. The impact of COVID-19 on pregnancy outcomes: a systematic review and meta-analysis. CMAJ Can Med Assoc J J Assoc Medicale Can. 2021 Apr 19;193(16):E540–8.

32. Child and infant mortality in England and Wales - Office for National Statistics [Internet]. [cited 2023 Jul 18]. Available from: https://www.ons.gov.uk/peoplepopulationandcommunity/birthsdeathsandmarriages/deaths/bulletins/childhoodinfantandperinatalmortalityinenglandandwales/2021

33. Stock SJ, Moore E, Calvert C, Carruthers J, Denny C, Donaghy J, et al. Pregnancy outcomes after SARS-CoV-2 infection in periods dominated by delta and omicron variants in Scotland: a population-based cohort study. Lancet Respir Med. 2022 Dec 1;10(12):1129–36.

