## Supplement for "Covid-19 vaccine safety in pregnancy, a nested case-control study in births from April 2021 to March 2022, England"

Figure 1: Data processing flow chart

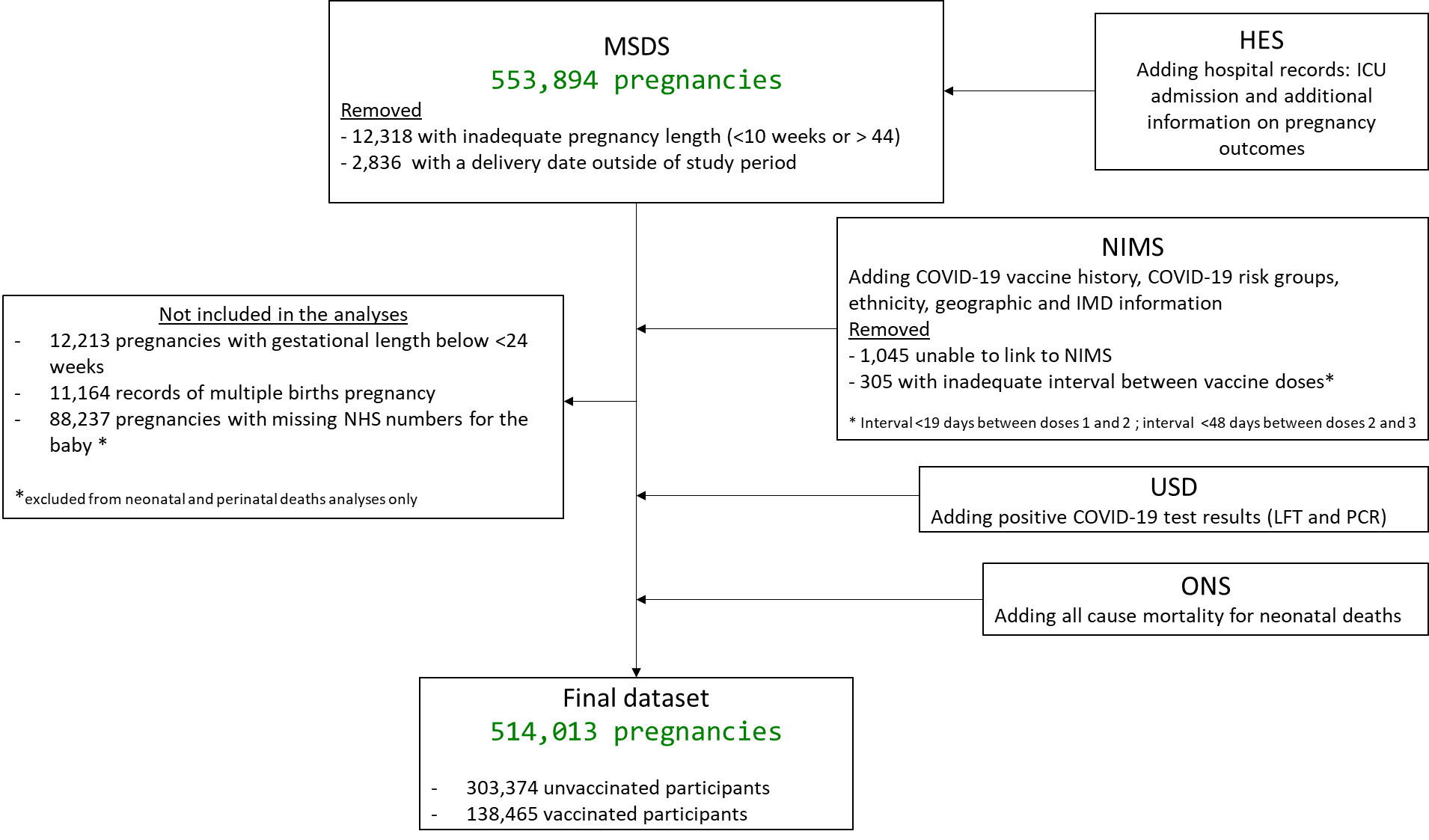

Supplement table 1: Case definition, matching criteria and index date for vaccine exposure used in the analyses

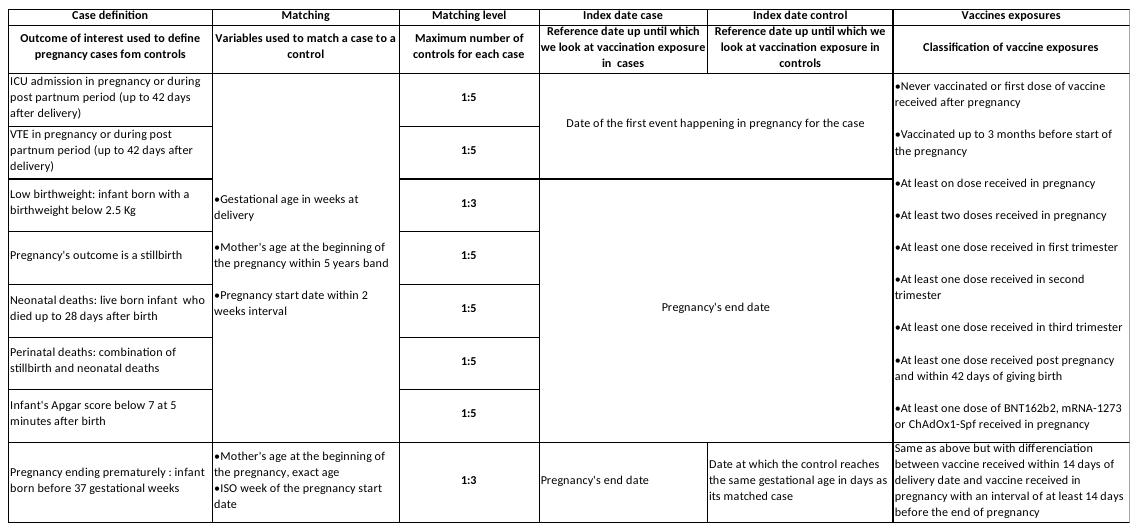

Supplement table 2: Number of controls matched to each case in the analyses

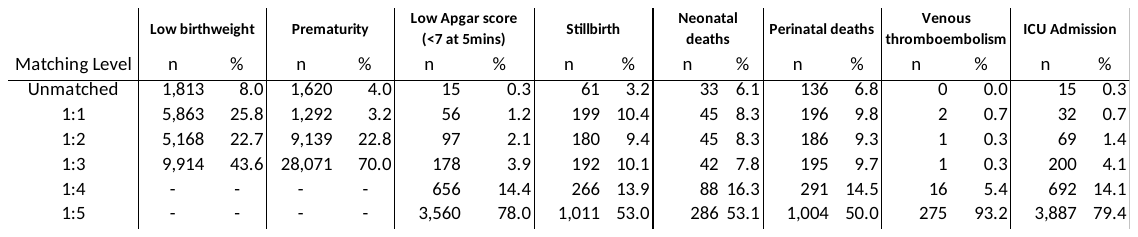

Supplement table 3: Demographic data on pregnant participants by outcomes status and P-Value

|  | **Low birthweight** | | | **Prematurity** | | | **Low Apgar score (<7 at 5mins)** | | |
| --- | --- | --- | --- | --- | --- | --- | --- | --- | --- |
| Risk factors / exposure | No | Yes | P-value | No | Yes | P-value | No | Yes | P-value |
| **Age groups in Years** | ***45,867*** | ***20,912*** |  | ***103,643*** | ***38,449*** |  | ***21,154*** | ***4,534*** |  |
| *18 to 24* | 8,665 (18.89) | 3,947 (18.87) |  | 16,050 (15.49) | 5,964 (15.51) |  | 3,674 (17.37) | 788 (17.38) |  |
| *25 to 29* | 12,737 (27.77) | 5,781 (27.64) | < 0.001** | 27,053 (26.10) | 9,896 (25.74) | < 0.001** | 6,091 (28.79) | 1,298 (28.63) | 0.011** |
| *30 to 34* | 14,385 (31.36) | 6,528 (31.22) |  | 33,789 (32.60) | 12,407 (32.27) |  | 6,896 (32.60) | 1,466 (32.33) |  |
| *35 to39* | 8,068 (17.59) | 3,722 (17.80) |  | 21,062 (20.32) | 7,889 (20.52) |  | 3,660 (17.30) | 786 (17.34) |  |
| *40 to 44* | 1,922 (4.19) | 885 (4.23) |  | 5,390 (5.20) | 2,128 (5.53) |  | 805 (3.81) | 187 (4.12) |  |
| *45 to 50* | 90 (0.20) | 49 (0.23) |  | 299 (0.29) | 165 (0.43) |  | 28 (0.13) | 9 (0.20) |  |
| **Ethnicity** | ***45,085*** | ***20,558*** |  | ***101,635*** | ***37,872*** |  | ***20,916*** | ***4,495*** |  |
| *White* | 33,439 (74.17) | 14,043 (68.31) | < 0.001** | 77,293 (76.05) | 27,423 (72.41) | < 0.001** | 15,775 (75.42) | 3,495 (77.75) | < 0.001** |
| *Mixed Multiple Ethnic groups* | 717 (1.59) | 370 (1.80) |  | 1,549 (1.52) | 633 (1.67) |  | 315 (1.51) | 73 (1.62) |  |
| *Black/Black British* | 2,740 (6.08) | 1,239 (6.03) |  | 5,397 (5.31) | 2,545 (6.72) |  | 1,135 (5.43) | 330 (7.34) |  |
| *Asian/Asian British* | 6,553 (14.53) | 4,259 (20.72) |  | 13,820 (13.60) | 5,925 (15.64) |  | 2,911 (13.92) | 489 (10.88) |  |
| *Any other ethnic groups* | 1,636 (3.63) | 647 (3.15) |  | 3,576 (3.52) | 1,346 (3.55) |  | 780 (3.73) | 108 (2.40) |  |
| **Region** | ***44,964*** | ***20,500*** |  | ***101,643*** | ***37,661*** |  | ***20,745*** | ***4,460*** |  |
| *East Midlands* | 3,676 (8.18) | 1,877 (9.16) |  | 8,313 (8.18) | 3,170 (8.42) |  | 1,665 (8.03) | 316 (7.09) |  |
| *East of England* | 5,126 (11.40) | 2,075 (10.12) |  | 12,232 (12.03) | 4,040 (10.73) |  | 2,403 (11.58) | 539 (12.09) |  |
| *London* | 8,105 (18.03) | 3,455 (16.85) | < 0.001** | 18,145 (17.85) | 6,747 (17.92) | 0.109** | 3,648 (17.58) | 596 (13.36) | < 0.001** |
| *North East* | 2,238 (4.98) | 824 (4.02) |  | 4,228 (4.16) | 1,717 (4.56) |  | 898 (4.33) | 301 (6.75) |  |
| *North West* | 6,308 (14.03) | 2,580 (12.59) |  | 12,995 (12.78) | 5,395 (14.33) |  | 2,782 (13.41) | 495 (11.10) |  |
| *South East* | 6,488 (14.43) | 2,891 (14.10) |  | 16,210 (15.95) | 5,409 (14.36) |  | 3,176 (15.31) | 732 (16.41) |  |
| *South West* | 3,807 (8.47) | 1,860 (9.07) |  | 9,050 (8.90) | 3,194 (8.48) |  | 1,859 (8.96) | 502 (11.26) |  |
| *West Midlands* | 5,170 (11.50) | 2,427 (11.84) |  | 10,792 (10.62) | 4,355 (11.56) |  | 2,278 (10.98) | 516 (11.57) |  |
| *Yorkshire and Humber* | 4,046 (9.00) | 2,511 (12.25) |  | 9,678 (9.52) | 3,634 (9.65) |  | 2,036 (9.81) | 463 (10.38) |  |
| **Index of Multiple Deprivation** | ***44,964*** | ***20,500*** |  | ***101,643*** | ***37,661*** |  | ***20,745*** | ***4,460*** |  |
| *1 Most Deprived* | 12,718 (28.28) | 6,497 (31.69) |  | 24,267 (23.87) | 11,227 (29.81) |  | 5,396 (26.01) | 1,203 (26.97) |  |
| *2* | 10,116 (22.50) | 4,907 (23.94) | < 0.001** | 21,683 (21.33) | 8,587 (22.80) | < 0.001** | 4,433 (21.37) | 993 (22.26) | < 0.001** |
| *3* | 8,390 (18.66) | 3,527 (17.20) |  | 19,761 (19.44) | 6,873 (18.25) |  | 4,046 (19.50) | 813 (18.23) |  |
| *4* | 7,305 (16.25) | 2,999 (14.63) |  | 18,546 (18.25) | 5,862 (15.57) |  | 3,640 (17.55) | 761 (17.06) |  |
| *5 Least Deprived* | 6,435 (14.31) | 2,570 (12.54) |  | 17,386 (17.10) | 5,112 (13.57) |  | 3,230 (15.57) | 690 (15.47) |  |
| **Complex social factors** | ***43,410*** | ***19,616*** |  | ***98,438*** | ***36,248*** |  | ***20,015*** | ***4,398*** |  |
| *No* | 36,384 (83.81) | 16,284 (83.01) | < 0.001* | 85,541 (86.90) | 30,216 (83.36) | < 0.001* | 17,203 (85.95) | 3,795 (86.29) | 0.431* |
| *Yes* | 7,026 (16.19) | 3,332 (16.99) |  | 12,897 (13.10) | 6,032 (16.64) |  | 2,812 (14.05) | 603 (13.71) |  |
| **Disability** | ***38,869*** | ***17,001*** |  | ***85,846*** | ***32,440*** |  | ***17,637*** | ***3,804*** |  |
| *No* | 37,218 (95.75) | 16,285 (95.79) |  | 82,770 (96.42) | 30,907 (95.27) |  | 16,975 (96.25) | 3,602 (94.69) |  |
| *Yes* | 1,651 (4.25) | 716 (4.21) | < 0.001* | 3,076 (3.58) | 1,533 (4.73) | < 0.001* | 662 (3.75) | 202 (5.31) | < 0.001* |
| **Parity** | ***45,941*** | ***20,945*** |  | ***103,783*** | ***38,502*** |  | ***21,208*** | ***4,547*** |  |
| *Nulliparous* | 17,281 (37.62) | 8,914 (42.56) |  | 41,405 (39.90) | 14,510 (37.69) |  | 8,525 (40.20) | 2,130 (46.84) |  |
| *Multiparous* | 28,660 (62.38) | 12,031 (57.44) | < 0.001* | 62,378 (60.10) | 23,992 (62.31) | < 0.001* | 12,683 (59.80) | 2,417 (53.16) | < 0.001* |
| **Gestational age at first appointment** | ***45,802*** | ***20,843*** |  | ***103,476*** | ***38,376*** |  | ***21,110*** | ***4,532*** |  |
| *< 12 weeks* | 38,282 (83.58) | 17,673 (84.79) |  | 91,467 (88.39) | 31,078 (80.98) |  | 18,253 (86.47) | 3,948 (87.11) |  |
| *1st Trimester* | 2,573 (5.62) | 1,139 (5.46) | < 0.001** | 4,930 (4.76) | 2,315 (6.03) | < 0.001** | 1,049 (4.97) | 230 (5.08) | 0.047** |
| *2nd Trimester* | 4,044 (8.83) | 1,676 (8.04) |  | 5,624 (5.44) | 4,120 (10.74) |  | 1,458 (6.91) | 280 (6.18) |  |
| *3rd Trimester* | 903 (1.97) | 355 (1.70) |  | 1,455 (1.41) | 863 (2.25) |  | 350 (1.66) | 74 (1.63) |  |
| **Previous adverse outcome** | ***45,941*** | ***20,945*** |  | ***103,783*** | ***38,502*** |  | ***21,208*** | ***4,547*** |  |
| *No* | 28,033 (61.02) | 12,378 (59.10) | < 0.001* | 65,375 (62.99) | 22,501 (58.44) | < 0.001* | 13,139 (61.95) | 2,749 (60.46) | 0.001* |
| *Yes* | 17,908 (38.98) | 8,567 (40.90) |  | 38,408 (37.01) | 16,001 (41.56) |  | 8,069 (38.05) | 1,798 (39.54) |  |
| **Comorbidities** | ***45,941*** | ***20,945*** |  | ***103,783*** | ***38,502*** |  | ***21,208*** | ***4,547*** |  |
| *No* | 43,203 (94.04) | 19,510 (93.15) | < 0.001* | 98,734 (95.14) | 35,905 (93.25) | < 0.001* | 20,055 (94.56) | 4,187 (92.08) | < 0.001* |
| *Yes* | 2,738 (5.96) | 1,435 (6.85) |  | 5,049 (4.86) | 2,597 (6.75) |  | 1,153 (5.44) | 360 (7.92) |  |
| **NHS worker** | ***45,824*** | ***20,909*** |  | ***103,483*** | ***38,412*** |  | ***21,157*** | ***4,538*** |  |
| *No* | 42,336 (92.39) | 19,495 (93.24) | < 0.001* | 95,414 (92.20) | 35,687 (92.91) | < 0.001* | 19,551 (92.41) | 4,199 (92.53) | 0.294* |
| *Yes* | 3,488 (7.61) | 1,414 (6.76) |  | 8,069 (7.80) | 2,725 (7.09) |  | 1,606 (7.59) | 339 (7.47) |  |
| **At risk of COVID-19 complications** | ***45,824*** | ***20,909*** |  | ***103,483*** | ***38,412*** |  | ***21,157*** | ***4,538*** |  |
| *No* | 34,377 (75.02) | 15,957 (76.32) |  | 83,604 (80.79) | 28,030 (72.97) |  | 16,752 (79.18) | 3,390 (74.70) |  |
| *Yes* | 11,447 (24.98) | 4,952 (23.68) | < 0.001* | 19,879 (19.21) | 10,382 (27.03) | < 0.001* | 4,405 (20.82) | 1,148 (25.30) | < 0.001* |
| **COVID-19 Infection in Pregnancy** | ***45,941*** | ***20,945*** |  | ***103,783*** | ***38,502*** |  | ***21,208*** | ***4,547*** |  |
| *No positive test in Pregnancy* | 39,310 (85.57) | 17,937 (85.64) | < 0.001* | 92,705 (89.33) | 33,400 (86.75) | < 0.001* | 18,735 (88.34) | 3,997 (87.90) | < 0.001* |
| *Tested positive in pregnancy before Index date* | 6,631 (14.43) | 3,008 (14.36) |  | 11,078 (10.67) | 5,102 (13.25) |  | 2,473 (11.66) | 550 (12.10) |  |

*P-Value from Chi2-test

** P-value from Kruskal–Wallis test (Chi2 with ties)

Supplement table 4 Demographic data on pregnant participants by outcomes status and P-Value

|  | **Venous thromboembolism** | | | **ICU Admission** | | |
| --- | --- | --- | --- | --- | --- | --- |
| Risk factors / exposure | No | Yes | P-value | No | Yes | P-value |
| **Age groups in Years** | ***1,446*** | ***295*** |  | ***22,941*** | ***4,872*** |  |
| *18 to 24* | 181 (12.52) | 38 (12.88) |  | 3,379 (14.73) | 710 (14.57) |  |
| *25 to 29* | 364 (25.17) | 74 (25.08) | 0.011** | 6,225 (27.13) | 1,315 (26.99) | < 0.001** |
| *30 to 34* | 496 (34.30) | 100 (33.90) |  | 7,624 (33.23) | 1,598 (32.80) |  |
| *35 to39* | 344 (23.79) | 70 (23.73) |  | 4,548 (19.82) | 972 (19.95) |  |
| *40 to 44* | 60 (4.15) | 12 (4.07) |  | 1,124 (4.90) | 263 (5.40) |  |
| *45 to 50* | 1 (0.07) | 1 (0.34) |  | 41 (0.18) | 14 (0.29) |  |
| **Ethnicity** | ***1,424*** | ***292*** |  | ***22,529*** | ***4,783*** |  |
| *White* | 1,104 (77.53) | 265 (90.75) | < 0.001** | 16,957 (75.27) | 2,402 (50.22) | < 0.001** |
| *Mixed Multiple Ethnic groups* | 15 (1.05) | 5 (1.71) |  | 351 (1.56) | 115 (2.40) |  |
| *Black/Black British* | 71 (4.99) | 6 (2.05) |  | 1,248 (5.54) | 598 (12.50) |  |
| *Asian/Asian British* | 193 (13.55) | 10 (3.42) |  | 3,181 (14.12) | 1,463 (30.59) |  |
| *Any other ethnic groups* | 41 (2.88) | 6 (2.05) |  | 792 (3.52) | 205 (4.29) |  |
| **Region** | ***1,415*** | ***288*** |  | ***22,510*** | ***4,785*** |  |
| *East Midlands* | 105 (7.42) | 14 (4.86) |  | 1,789 (7.95) | 328 (6.85) |  |
| *East of England* | 165 (11.66) | 15 (5.21) |  | 2,633 (11.70) | 203 (4.24) |  |
| *London* | 258 (18.23) | 36 (12.50) | < 0.001** | 3,926 (17.44) | 1,706 (35.65) | < 0.001** |
| *North East* | 57 (4.03) | 12 (4.17) |  | 989 (4.39) | 71 (1.48) |  |
| *North West* | 172 (12.16) | 39 (13.54) |  | 3,070 (13.64) | 825 (17.24) |  |
| *South East* | 237 (16.75) | 45 (15.63) |  | 3,466 (15.40) | 280 (5.85) |  |
| *South West* | 148 (10.46) | 4 (1.39) |  | 2,001 (8.89) | 110 (2.30) |  |
| *West Midlands* | 129 (9.12) | 86 (29.86) |  | 2,444 (10.86) | 1,122 (23.45) |  |
| *Yorkshire and Humber* | 144 (10.18) | 37 (12.85) |  | 2,192 (9.74) | 140 (2.93) |  |
| **Index of Multiple Deprivation** | ***1,415*** | ***288*** |  | ***22,510*** | ***4,785*** |  |
| *1 Most Deprived* | 312 (22.05) | 88 (30.56) |  | 5,753 (25.56) | 1,706 (35.65) |  |
| *2* | 326 (23.04) | 67 (23.26) | 0.010** | 4,849 (21.54) | 1,422 (29.72) | < 0.001** |
| *3* | 263 (18.59) | 46 (15.97) |  | 4,241 (18.84) | 757 (15.82) |  |
| *4* | 269 (19.01) | 46 (15.97) |  | 4,078 (18.12) | 501 (10.47) |  |
| *5 Least Deprived* | 245 (17.31) | 41 (14.24) |  | 3,589 (15.94) | 399 (8.34) |  |
| **Complex social factors** | ***1,380*** | ***293*** |  | ***21,595*** | ***4,674*** |  |
| *No* | 1,209 (87.61) | 255 (87.03) | 0.862* | 18,564 (85.96) | 3,974 (85.02) | < 0.001* |
| *Yes* | 171 (12.39) | 38 (12.97) |  | 3,031 (14.04) | 700 (14.98) |  |
| **Disability** | ***1,204*** | ***289*** |  | ***19,011*** | ***4,444*** |  |
| *No* | 1,152 (95.68) | 253 (87.54) |  | 18,258 (96.04) | 4,195 (94.40) |  |
| *Yes* | 52 (4.32) | 36 (12.46) | < 0.001* | 753 (3.96) | 249 (5.60) | < 0.001* |
| **Parity** | ***1,446*** | ***295*** |  | ***22,973*** | ***4,880*** |  |
| *Nulliparous* | 565 (39.07) | 73 (24.75) |  | 8,848 (38.51) | 1,982 (40.61) |  |
| *Multiparous* | 881 (60.93) | 222 (75.25) | < 0.001* | 14,125 (61.49) | 2,898 (59.39) | 0.420* |
| **Gestational age at first appointment** | ***1,445*** | ***295*** |  | ***22,903*** | ***4,814*** |  |
| *< 12 weeks* | 1,243 (86.02) | 254 (86.10) |  | 19,753 (86.25) | 3,950 (82.05) |  |
| *1st Trimester* | 80 (5.54) | 20 (6.78) | 0.3559 | 1,168 (5.10) | 315 (6.54) | < 0.001** |
| *2nd Trimester* | 100 (6.92) | 17 (5.76) |  | 1,589 (6.94) | 462 (9.60) |  |
| *3rd Trimester* | 22 (1.52) | 4 (1.36) |  | 393 (1.72) | 87 (1.81) |  |
| **Previous adverse outcome** | ***1,446*** | ***295*** |  | ***22,973*** | ***4,880*** |  |
| *No* | 883 (61.07) | 144 (48.81) | < 0.001* | 14,098 (61.37) | 2,801 (57.40) | < 0.001* |
| *Yes* | 563 (38.93) | 151 (51.19) |  | 8,875 (38.63) | 2,079 (42.60) |  |
| **Comorbidities** | ***1,446*** | ***295*** |  | ***22,973*** | ***4,880*** |  |
| *No* | 883 (61.07) | 144 (48.81) | < 0.001* | 21,712 (94.51) | 4,474 (91.68) | < 0.001* |
| *Yes* | 563 (38.93) | 151 (51.19) |  | 1,261 (5.49) | 406 (8.32) |  |
| **NHS worker** | ***1,444*** | ***295*** |  | ***22,916*** | ***4,868*** |  |
| *No* | 1,315 (91.07) | 280 (94.92) | 0.076* | 21,120 (92.16) | 4,524 (92.93) | 0.037* |
| *Yes* | 129 (8.93) | 15 (5.08) |  | 1,796 (7.84) | 344 (7.07) |  |
| **At risk of COVID-19 complications** | ***1,444*** | ***295*** |  | ***22,916*** | ***4,868*** |  |
| *No* | 1,167 (80.82) | 108 (36.61) |  | 18,037 (78.71) | 3,261 (66.99) |  |
| *Yes* | 277 (19.18) | 187 (63.39) | < 0.001* | 4,879 (21.29) | 1,607 (33.01) | < 0.001* |
| **COVID-19 Infection in Pregnancy** | ***1,446*** | ***295*** |  | ***22,973*** | ***4,880*** |  |
| *No positive test in Pregnancy* | 316 (86.81) | 65 (84.42) | < 0.001* | 20,266 (88.22) | 3,902 (79.96) | < 0.001* |
| *Tested positive in pregnancy before Index date* | 48 (13.19) | 12 (15.58) |  | 2,707 (11.78) | 978 (20.04) |  |

*P-Value from Chi2-test

** P-value from Kruskal–Wallis test (Chi2 with ties)

Supplement table 5 Demographic data on pregnant participants by outcomes status and P-Value

|  | **Venous thromboembolism** | | | **ICU Admission** | | |
| --- | --- | --- | --- | --- | --- | --- |
| Risk factors / exposure | No | Yes | P-value | No | Yes | P-value |
| **Age groups in Years** | ***1,446*** | ***295*** |  | ***22,941*** | ***4,872*** |  |
| *18 to 24* | 181 (12.52) | 38 (12.88) |  | 3,379 (14.73) | 710 (14.57) |  |
| *25 to 29* | 364 (25.17) | 74 (25.08) | 0.011** | 6,225 (27.13) | 1,315 (26.99) | < 0.001** |
| *30 to 34* | 496 (34.30) | 100 (33.90) |  | 7,624 (33.23) | 1,598 (32.80) |  |
| *35 to39* | 344 (23.79) | 70 (23.73) |  | 4,548 (19.82) | 972 (19.95) |  |
| *40 to 44* | 60 (4.15) | 12 (4.07) |  | 1,124 (4.90) | 263 (5.40) |  |
| *45 to 50* | 1 (0.07) | 1 (0.34) |  | 41 (0.18) | 14 (0.29) |  |
| **Ethnicity** | ***1,424*** | ***292*** |  | ***22,529*** | ***4,783*** |  |
| *White* | 1,104 (77.53) | 265 (90.75) | < 0.001** | 16,957 (75.27) | 2,402 (50.22) | < 0.001** |
| *Mixed Multiple Ethnic groups* | 15 (1.05) | 5 (1.71) |  | 351 (1.56) | 115 (2.40) |  |
| *Black/Black British* | 71 (4.99) | 6 (2.05) |  | 1,248 (5.54) | 598 (12.50) |  |
| *Asian/Asian British* | 193 (13.55) | 10 (3.42) |  | 3,181 (14.12) | 1,463 (30.59) |  |
| *Any other ethnic groups* | 41 (2.88) | 6 (2.05) |  | 792 (3.52) | 205 (4.29) |  |
| **Region** | ***1,415*** | ***288*** |  | ***22,510*** | ***4,785*** |  |
| *East Midlands* | 105 (7.42) | 14 (4.86) |  | 1,789 (7.95) | 328 (6.85) |  |
| *East of England* | 165 (11.66) | 15 (5.21) |  | 2,633 (11.70) | 203 (4.24) |  |
| *London* | 258 (18.23) | 36 (12.50) | < 0.001** | 3,926 (17.44) | 1,706 (35.65) | < 0.001** |
| *North East* | 57 (4.03) | 12 (4.17) |  | 989 (4.39) | 71 (1.48) |  |
| *North West* | 172 (12.16) | 39 (13.54) |  | 3,070 (13.64) | 825 (17.24) |  |
| *South East* | 237 (16.75) | 45 (15.63) |  | 3,466 (15.40) | 280 (5.85) |  |
| *South West* | 148 (10.46) | 4 (1.39) |  | 2,001 (8.89) | 110 (2.30) |  |
| *West Midlands* | 129 (9.12) | 86 (29.86) |  | 2,444 (10.86) | 1,122 (23.45) |  |
| *Yorkshire and Humber* | 144 (10.18) | 37 (12.85) |  | 2,192 (9.74) | 140 (2.93) |  |
| **Index of Multiple Deprivation** | ***1,415*** | ***288*** |  | ***22,510*** | ***4,785*** |  |
| *1 Most Deprived* | 312 (22.05) | 88 (30.56) |  | 5,753 (25.56) | 1,706 (35.65) |  |
| *2* | 326 (23.04) | 67 (23.26) | 0.010** | 4,849 (21.54) | 1,422 (29.72) | < 0.001** |
| *3* | 263 (18.59) | 46 (15.97) |  | 4,241 (18.84) | 757 (15.82) |  |
| *4* | 269 (19.01) | 46 (15.97) |  | 4,078 (18.12) | 501 (10.47) |  |
| *5 Least Deprived* | 245 (17.31) | 41 (14.24) |  | 3,589 (15.94) | 399 (8.34) |  |
| **Complex social factors** | ***1,380*** | ***293*** |  | ***21,595*** | ***4,674*** |  |
| *No* | 1,209 (87.61) | 255 (87.03) | 0.862* | 18,564 (85.96) | 3,974 (85.02) | < 0.001* |
| *Yes* | 171 (12.39) | 38 (12.97) |  | 3,031 (14.04) | 700 (14.98) |  |
| **Disability** | ***1,204*** | ***289*** |  | ***19,011*** | ***4,444*** |  |
| *No* | 1,152 (95.68) | 253 (87.54) |  | 18,258 (96.04) | 4,195 (94.40) |  |
| *Yes* | 52 (4.32) | 36 (12.46) | < 0.001* | 753 (3.96) | 249 (5.60) | < 0.001* |
| **Parity** | ***1,446*** | ***295*** |  | ***22,973*** | ***4,880*** |  |
| *Nulliparous* | 565 (39.07) | 73 (24.75) |  | 8,848 (38.51) | 1,982 (40.61) |  |
| *Multiparous* | 881 (60.93) | 222 (75.25) | < 0.001* | 14,125 (61.49) | 2,898 (59.39) | 0.420* |
| **Gestational age at first appointment** | ***1,445*** | ***295*** |  | ***22,903*** | ***4,814*** |  |
| *< 12 weeks* | 1,243 (86.02) | 254 (86.10) |  | 19,753 (86.25) | 3,950 (82.05) |  |
| *1st Trimester* | 80 (5.54) | 20 (6.78) | 0.3559 | 1,168 (5.10) | 315 (6.54) | < 0.001** |
| *2nd Trimester* | 100 (6.92) | 17 (5.76) |  | 1,589 (6.94) | 462 (9.60) |  |
| *3rd Trimester* | 22 (1.52) | 4 (1.36) |  | 393 (1.72) | 87 (1.81) |  |
| **Previous adverse outcome** | ***1,446*** | ***295*** |  | ***22,973*** | ***4,880*** |  |
| *No* | 883 (61.07) | 144 (48.81) | < 0.001* | 14,098 (61.37) | 2,801 (57.40) | < 0.001* |
| *Yes* | 563 (38.93) | 151 (51.19) |  | 8,875 (38.63) | 2,079 (42.60) |  |
| **Comorbidities** | ***1,446*** | ***295*** |  | ***22,973*** | ***4,880*** |  |
| *No* | 883 (61.07) | 144 (48.81) | < 0.001* | 21,712 (94.51) | 4,474 (91.68) | < 0.001* |
| *Yes* | 563 (38.93) | 151 (51.19) |  | 1,261 (5.49) | 406 (8.32) |  |
| **NHS worker** | ***1,444*** | ***295*** |  | ***22,916*** | ***4,868*** |  |
| *No* | 1,315 (91.07) | 280 (94.92) | 0.076* | 21,120 (92.16) | 4,524 (92.93) | 0.037* |
| *Yes* | 129 (8.93) | 15 (5.08) |  | 1,796 (7.84) | 344 (7.07) |  |
| **At risk of COVID-19 complications** | ***1,444*** | ***295*** |  | ***22,916*** | ***4,868*** |  |
| *No* | 1,167 (80.82) | 108 (36.61) |  | 18,037 (78.71) | 3,261 (66.99) |  |
| *Yes* | 277 (19.18) | 187 (63.39) | < 0.001* | 4,879 (21.29) | 1,607 (33.01) | < 0.001* |
| **COVID-19 Infection in Pregnancy** | ***1,446*** | ***295*** |  | ***22,973*** | ***4,880*** |  |
| *No positive test in Pregnancy* | 316 (86.81) | 65 (84.42) | < 0.001* | 20,266 (88.22) | 3,902 (79.96) | < 0.001* |
| *Tested positive in pregnancy before Index date* | 48 (13.19) | 12 (15.58) |  | 2,707 (11.78) | 978 (20.04) |  |

*P-Value from Chi2-test

** P-value from Kruskal–Wallis test (Chi2 with ties)

Supplement table 6: SNOMED code list of comorbidities and health related risk factors in pregnancy

| **SNOMED code** | **Reference** | **Categorisation** |
| --- | --- | --- |
| 234446004 | vWD - Congenital von Willebrand's disease | Bleeding disorder |
| 128105004 | vWD - von Willebrand's disease | Bleeding disorder |
| 70142008 | Atrial septal defect (disorder) | Cardiac |
| 275516004 | Cardiomegaly - hypertensive (disorder) | Cardiac |
| 56265001 | Heart diseases | Cardiac |
| 161625008 | History of cardiac surgery (situation) | cardiac |
| 48724000 | MR - Mitral regurgitation | Cardiac |
| 10759031000119106 | Pre-existing hypertensive heart disease in pregnancy | Cardiac |
| 62377009 | Puerperal cardiomyopathy | Cardiac |
| 56786000 | PVS - Pulmonary valve stenosis | Cardiac |
| 111287006 | Tricuspid valve regurgitation, NOS | cardiac |
| 368009 | Valvular heart disease, NOS | Cardiac |
| 74390002 | WPW - Wolff-Parkinson-White syndrome | Cardiac |
| 720521008 | Autosomal dominant macrothrombocytopenia (disorder) | Coagulation |
| 307091009 | Factor V Leiden mutation (disorder) | Coagulation |
| 307116001 | Heterozygous Factor V Leiden mutation (disorder) | Coagulation |
| 724637001 | Isolated thrombocytopenia (disorder) | Coagulation |
| 76407009 | Protein C deficiency disease (disorder) | Coagulation |
| 1563006 | Protein S deficiency disease (disorder) | Coagulation |
| 302215000 | Thrombocytopenic disorder (disorder) | Coagulation |
| 72866009 | VV - Varicose veins of leg | Coagulation risk |
| 73211009 | Diabetes mellitus (disorder) | Diabetes |
| 426705001 | Diabetes mellitus co-occurrent and due to cystic fibrosis (disorder) | Diabetes type 1 |
| 427089005 | Diabetes mellitus due to cystic fibrosis (disorder) | Diabetes type 1 |
| 46635009 | Diabetes mellitus type 1 (disorder) | Diabetes type 1 |
| 46635009 | Diabetes mellitus type 1 (disorder) | Diabetes type 1 |
| 44054006 | Diabetes mellitus type 2 (disorder) | Diabetes Type 2 |
| 44054006 | Diabetes mellitus type 2 (disorder) | Diabetes type 2 |
| 81531005 | Diabetes mellitus type 2 in obese (disorder) | Diabetes Type 2 |
| 164971000119101 | Type 2 diabetes mellitus controlled by diet (finding) | Diabetes Type 2 |
| 24481000000101 | Type 2 diabetes on diet only (finding) | Diabetes Type 2 |
| 313436004 | Type II diabetes mellitus without complication (disorder) | Diabetes Type 2 |
| 199223000 | Diabetes mellitus during pregnancy, childbirth and the puerperium (disorder) | Diabetes, GDM |
| 11687002 | Gestational diabetes mellitus (disorder) | Diabetes, GDM |
| 40801000119106 | Gestational diabetes mellitus complicating pregnancy (disorder) | Diabetes, GDM |
| 40791000119105 | Postpartum gestational diabetes mellitus (disorder) | Diabetes, GDM |
| 429094000 | Dietary advice for type I diabetes | Diabetes, Type 1 |
| 444073006 | Type I diabetes mellitus uncontrolled (finding) | Diabetes, Type 1 |
| 73211009 | Diabetes mellitus (disorder) | Diabetes, unspecified |
| 170763003 | Diabetic - good control (finding) | Diabetes, unspecified |
| 170745003 | Diabetic on diet only (finding) | Diabetes, unspecified |
| 170747006 | Diabetic on insulin (finding) | Diabetes, unspecified |
| 137421000119106 | Graves' disease in remission (disorder) | Endocrine |
| 428165003 | Hypothyroidism in pregnancy (disorder) | Endocrine |
| 253011004 | Macroprolactinoma (disorder) | Endocrine |
| 253010003 | Microprolactinoma (disorder) | Endocrine |
| 254956000 | Pituitary adenoma (disorder) | Endocrine |
| 399244003 | Pituitary disease | Endocrine |
| 10649000 | Pituitary hyperfunction, NOS | Endocrine |
| 254965007 | Pituitary macroadenoma (disorder) | Endocrine |
| 254963000 | Pituitary microadenoma (disorder) | Endocrine |
| 36348003 | Primary hyperparathyroidism (disorder) | Endocrine |
| 82119001 | Thyroiditis, NOS | Endocrine |
| 237662005 | Hyperprolactinemia (disorder) | Endocrine, other |
| 352818000 | Tonic-clonic epilepsy (disorder) | Epilepsy |
| 1201005 | Benign essential hypertension (disorder) | Hypertension |
| 843841000000109 | Severe hypertension (National Institute for Health and Clinical Excellence 2011) (disorder) | Hypertension |
| 71874008 | Benign essential hypertension complicating or reason for care during childbirth | Hypertension - eclampsia/pre-eclampsia |
| 8218002 | Chronic hypertension complicating or reason for care during childbirth | Hypertension - eclampsia/pre-eclampsia |
| 8762007 | Chronic hypertension in obstetric context (disorder) | Hypertension - eclampsia/pre-eclampsia |
| 198992004 | Eclampsia in pregnancy (disorder) | Hypertension - eclampsia/pre-eclampsia |
| 426634003 | Eclamptic seizure (finding) | Hypertension - eclampsia/pre-eclampsia |
| 41114007 | PET - Mild pre-eclamptic toxemia | Hypertension - eclampsia/pre-eclampsia |
| 67359005 | Pre-eclampsia added to pre-existing hypertension (disorder) | Hypertension - eclampsia/pre-eclampsia |
| 765182005 | Pre-eclampsia in puerperium (disorder) | Hypertension - eclampsia/pre-eclampsia |
| 398254007 | Proteinuric hypertension of pregnancy | Hypertension - eclampsia/pre-eclampsia |
| 46764007 | Severe proteinuric hypertension of pregnancy (disorder) | Hypertension - eclampsia/pre-eclampsia |
| 59621000 | Essential hypertension (disorder) | Hypertension (pre-existing) |
| 72022006 | Essential hypertension in obstetric context, NOS | Hypertension (pre-existing) |
| 169465000 | Hypertension caused by oral contraceptive pill (disorder) | Hypertension, chronic |
| 40521000119100 | Postpartum pregnancy-induced hypertension (disorder) | Hypertension, PIH |
| 82771000119102 | Hypertension complicating pregnancy (disorder) | Hypertension, unclassified |
| 198941007 | Hypertension complicating pregnancy, childbirth and the puerperium (disorder) | Hypertension, unclassified |
| 367390009 | Hypertension without albuminuria AND without oedema in the obstetric context | Hypertension, unclassified |
| 38341003 | Hypertensive disorder, systemic arterial (disorder) | Hypertension, unclassified |
| 48194001 | Unspecified hypertension complicating pregnancy, childbirth or puerperium | Hypertension, unspecified |
| 353295004 | Graves' disease (disorder) | Hyperthyroid |
| 128477000 | Abscess (disorder) | Infection |
| 13802001 | Abscess of axilla (disorder) | Infection |
| 69430001 | Abscess of vulva (disorder) | Infection |
| 10850741000119108 | Accidental needle stick injury (disorder) | Infection |
| 85189001 | Acute appendicitis, NOS | Infection |
| 85189001 | Acute appendicitis, NOS | Infection |
| 40331000119107 | Acute cholecystitis due to biliary calculus (disorder) | Infection |
| 704175009 | Acute chorioamnionitis (disorder) | Infection |
| 67667007 | Acute endometritis (disorder) | Infection |
| 64994000 | Acute fulminating appendicitis with perforation AND peritonitis (disorder) | Infection |
| 69776003 | Acute gastroenteritis, NOS | Infection |
| 82789004 | Acute mastitis (disorder) | Infection |
| 36689008 | APN - Acute pyelonephritis | Infection |
| 74400008 | Appendicitis, NOS | Infection |
| 53084003 | Bacterial pneumonia (disorder) | Infection |
| 10001005 | Bacterial septicaemia, NOS | Infection |
| 312124009 | Bacterial UTI (urinary tract infection) | Infection |
| 198108005 | Breast infection (disorder) | Infection |
| 396285007 | Bronchopneumonia (disorder) | Infection |
| 128045006 | Cellulitis (disorder) | Infection |
| 385627004 | Cellulitis (morphologic abnormality) | Infection |
| 409799006 | Extended spectrum beta-lactamase producing bacteria (organism) | infection |
| 726500000 | Extended spectrum beta-lactamase producing Enterobacteriaceae (organism) | infection |
| 409800005 | Extended spectrum beta-lactamse producing Escherichia coli (organism) | infection |
| 25374005 | GE - Gastroenteritis | Infection |
| 10756101000119107 | Group B streptococcus infection in mother complicating childbirth (disorder) | Infection |
| 237339000 | Infection - perineal wound (disorder) | Infection |
| 40733004 | Infective disorder | Infection |
| 40468003 | Infective hepatitis | Infection |
| 174041007 | Laparoscopic emergency appendicectomy (procedure) | Infection |
| 11999007 | Latent tuberculosis | Infection |
| 50417007 | Lower respiratory tract infection (disorder) | Infection |
| 431737008 | Lower urinary tract infection of sudden onset AND/OR short duration | Infection |
| 95883001 | Meningitis, bacterial | Infection |
| 7180009 | Meningitis, NOS | Infection |
| 198130006 | PID - pelvic inflammatory disease | Infection |
| 431709001 | Pilonidal abscess of natal cleft (disorder) | Infection |
| 51169003 | Pneumococcal meningitis (disorder) | Infection |
| 233604007 | Pneumonia (disorder) | Infection |
| 882784691000119100 | Pneumonia caused by severe acute respiratory syndrome coronavirus 2 (disorder) | Infection |
| 441942006 | Pneumonia due to infection caused by Streptococcus pyogenes | Infection |
| 1111341000000102 | Presence of human immunodeficiency virus 1 antibody in serum (observable entity) | Infection |
| 700038005 | Puerperal mastitis | Infection |
| 237348005 | Puerperal pyrexia (disorder) | Infection |
| 2858002 | Puerperal septicaemia (disorder) | Infection |
| 16271000119108 | Pyelonephritis in pregnancy (disorder) | Infection |
| 45816000 | Pyelonephritis, NOS | Infection |
| 197927001 | Recurrent UTI - urinary tract infection | Infection |
| 12463005 | Septic gastroenteritis, NOS | Infection |
| 1125006 | Septicaemia during labor (disorder) | Infection |
| 1036671000000106 | Severe sepsis (disorder) | Infection |
| 43492007 | Streptococcus, group B | Infection |
| 91302008 | Systemic infection, NOS | Infection |
| 238149007 | Systemic inflammatory response syndrome (disorder) | Infection |
| 187192000 | Toxoplasmosis (disorder) | Infection |
| 415760001 | Tuberculosis status (observable entity) | Infection |
| 56717001 | Tuberculosis, NOS | Infection |
| 54150009 | Upper respiratory infection (disorder) | Infection |
| 431308006 | Upper urinary tract infection of sudden onset AND/OR short duration | Infection |
| 700372006 | Urinary tract infection associated with catheter (disorder) | Infection |
| 431309003 | Urinary tract infection of sudden onset AND/OR short duration | Infection |
| 167570003 | Urine culture - E. coli (finding) | Infection |
| 301775005 | Uterine infection | Infection |
| 4009004 | UTI - Lower urinary tract infection | Infection |
| 68566005 | UTI - Urinary tract infection | Infection |
| 307534009 | UTI - urinary tract infection in pregnancy | Infection |
| 38907003 | Varicella, NOS | Infection |
| 309465005 | Varicella-zoster virus infection (disorder) | Infection |
| 25102003 | Viral (infectious) hepatitis A | Infection |
| 66071002 | Viral hepatitis type B (disorder) | Infection |
| 50711007 | Viral hepatitis, non-A, non-B | Infection |
| 34014006 | Viral infectious disease, NOS | Infection |
| 75570004 | Viral pneumonia (disorder) | Infection |
| 51209006 | Viral tonsillitis (disorder) | Infection |
| 413643004 | Group A Strep | Infection |
| 407451003 | Herpes simplex type 1 infection (disorder) | Infection |
| 423391007 | Herpes simplex type 2 genital infection | Infection |
| 1631000119103 | Group B Streptococcus carrier (finding) | Infection -GBS |
| 120001000119107 | Group B streptococcus carrier complicating pregnancy (disorder) | Infection -GBS |
| 426933007 | Group B streptococcus infection | Infection -GBS |
| 371012000 | Acute lymphoblastic leukaemia, transitional pre-B-cell (disorder) | Other medical |
| 403486000 | Acute systemic lupus erythematosus (disorder) | Other medical |
| 128600008 | Acute ulcerative colitis (disorder) | Other medical |
| 363732003 | Addison's disease (disorder) | Other medical |
| 197456007 | AP - Acute pancreatitis | Other medical |
| 26843008 | APS - Antiphospholipid syndrome | Other medical |
| 274135002 | Arthritis/arthrosis (disorder) | Other medical |
| 85828009 | Autoimmune disorder, NOS | Other medical |
| 237519003 | Autoimmune hypothyroidism (disorder) | Other medical |
| 310701003 | Behcet's syndrome (disorder) | Other medical |
| 15523002 | Benign focal epilepsy of childhood (disorder) | Other medical |
| 269175006 | Beta trait thalassaemia | Other medical |
| 79592006 | beta^+^ Thalassemia, NOS | Other medical |
| 412734009 | Breast cancer 1, early onset gene mutation positive | Other medical |
| 412738007 | Breast cancer 2, early onset gene mutation positive | Other medical |
| 12295008 | Bronchiectasis (disorder) | Other medical |
| 78004001 | Bulimia nervosa (disorder) | Other medical |
| 230690007 | Cerebrovascular accident (disorder) | Other medical |
| 253184003 | Chiari malformation (disorder) | Other medical |
| 253185002 | Chiari malformation type I (disorder) | Other medical |
| 6382002 | Chronic inflammatory small bowel disease (disorder) | Other medical |
| 92564006 | CIS - Carcinoma in situ of cervix | Other medical |
| 83470009 | Ehlers-Danlos syndrome, type 1 (disorder) | Other medical |
| 84757009 | Epilepsy (disorder) | Other medical |
| 100941000119100 | Epilepsy in pregnancy | Other medical |
| 608844008 | History of inflammatory bowel disease (situation) | other medical |
| 74728003 | Hypopituitarism (disorder) | other medical |
| 191306005 | IgA vasculitis | Other medical |
| 277637000 | Large cell anaplastic lymphoma (disorder) | Other medical |
| 277569004 | Large granular lymphocytic leukaemia (disorder) | Other medical |
| 193022009 | Localization-related(focal)(partial)idiopathic epilepsy and epileptic syndromes with seizures of localized onset (disorder) | Other medical |
| 200936003 | Lupus erythematosus (disorder) | Other medical |
| 14537002 | Malignant lymphoma, Hodgkin's | Other medical |
| 129597002 | Moderate hyperemesis gravidarum (disorder) | Other medical |
| 230445007 | Nocturnal epilepsy (disorder) | Other medical |
| 9014002 | Psoriasis, NOS | Other medical |
| 156370009 | Psoriatic arthritis (disorder) | Other medical |
| 33339001 | Psoriatic arthropathy | Other medical |
| 195295006 | Raynaud's disease (disorder) | Other medical |
| 426373005 | Relapsing remitting multiple sclerosis (disorder) | Other medical |
| 90708001 | Renal disorder, NOS | Other medical |
| 123803006 | Rheumatic arteritis (disorder) | Other medical |
| 399923009 | Rheumatoid arteritis (disorder) | Other medical |
| 69896004 | Rheumatoid arthritis (disorder) | Other medical |
| 129635004 | Secondary Meig's syndrome | Other medical |
| 370221004 | Severe asthma (disorder) | Other medical |
| 83901003 | Sicca (Sjogren's) syndrome | Other medical |
| 16402000 | Sickle cells present | Other medical |
| 49938009 | Sickling | Other medical |
| 19267009 | SLE inhibitor syndrome | Other medical |
| 32232003 | Spina bifida of cervical region (disorder) | Other medical |
| 87163000 | Subacute leukaemia [obs] | Other medical |
| 21454007 | Subarachnoid intracranial haemorrhage (disorder) | Other medical |
| 54823002 | Subclinical hypothyroidism (disorder) | Other medical |
| 2477008 | Superficial thrombophlebitis, NOS | Other medical |
| 71831005 | Symptomatic generalized epilepsy (disorder) | Other medical |
| 68815009 | Systemic lupus erythematosus glomerulonephritis syndrome (disorder) | Other medical |
| 73286009 | Systemic lupus erythematosus glomerulonephritis syndrome, World Health Organization class I (disorder) | Other medical |
| 19442009 | Thalassemia trait, NOS | Other medical |
| 40108008 | Thalassemia, NOS | Other medical |
| 36472007 | Thalassemia-haemoglobin S disease | Other medical |
| 78514002 | Thigh pain (finding) | Other medical |
| 64766004 | Ulcerative colitis, NOS | Other medical |
| 52231000 | Ulcerative proctitis | Other medical |
| 86049000 | Unclassified tumour, malignant | Other medical |
| 724601006 | Vasculitis caused by antineutrophil cytoplasmic antibody (disorder) | Other medical |
| 31996006 | Vasculitis, NOS | Other medical |
| 733858005 | Acute severe refractory exacerbation of asthma (disorder) | Other medical (asthma) |
| 10754921000119106 | Hyperthyroidism in childbirth (disorder) | Thyroid disorder |
| 72271000119100 | Hyperthyroidism in pregnancy (disorder) | Thyroid disorder |
| 34486009 | Hyperthyroidism, NOS | Thyroid disorder |
| 40930008 | Hypothyroidism (disorder) | Thyroid disorder |
| 341861000000109 | Hypothyroidism annual review (regime/therapy) | Thyroid disorder |
| 237520009 | Hypothyroidism due to Hashimoto's thyroiditis (disorder) | Thyroid disorder |
| 205101000000104 | Hypothyroidism review (regime/therapy) | Thyroid disorder |
| 264580006 | Thyroid dysfunction (disorder) | Thyroid disorder |

Supplement table 7: ICD-10 code list of comorbidities and health related risk factors in pregnancy

| **ICD-10 code** | **Reference** | **Categorisation** |
| --- | --- | --- |
| D680 | Von Willebrand disease | Bleeding disorder |
| I071 | Tricuspid insufficiency | Cardiac |
| I099 | Rheumatic heart disease, unspecified | Cardiac |
| I350 | Aortic (valve) stenosis | Cardiac |
| I351 | Aortic (valve) insufficiency | Cardiac |
| I352 | Aortic (valve) stenosis with insufficiency | Cardiac |
| I370 | Pulmonary valve stenosis | Cardiac |
| I371 | Pulmonary valve insufficiency | Cardiac |
| I429 | Cardiomyopathy, unspecified | Cardiac |
| I489 | Atrial fibrillation and atrial flutter, unspecified | Cardiac |
| I499 | Cardiac arrhythmia, unspecified | Cardiac |
| I509 | Heart failure, unspecified | cardiac |
| I517 | Cardiomegaly | Cardiac |
| J81X | Pulmonary oedema | Cardiac |
| O101 | Pre-existing hypertensive heart disease complicating pregnancy, childbirth and the puerperium | Cardiac |
| O903 | Cardiomyopathy in the puerperium | Cardiac |
| Q210 | Ventricular septal defect | Cardiac |
| Q211 | Atrial septal defect | Cardiac |
| Q213 | Tetralogy of Fallot | Cardiac |
| Q220 | Pulmonary valve atresia | Cardiac |
| Q234 | Hypoplastic left heart syndrome | cardiac |
| Z941 | Heart transplant status | cardiac |
| D691 | Qualitative platelet defects | Clotting |
| D695 | Secondary thrombocytopenia | Clotting |
| D696 | Thrombocytopenia, unspecified | Clotting |
| D751 | Secondary polycythaemia | Clotting |
| D685 | Primary Thrombophilia | Coagulation |
| E103 | Type 1 diabetes mellitus: With ophthalmic complications | Diabetes Type 1 |
| E109 | Type 1 diabetes mellitus: Without complications | Diabetes Type 2 |
| O241 | Diabetes mellitus in pregnancy: Pre-existing type 2 diabetes mellitus | Diabetes Type 2 |
| E113 | Type 2 diabetes mellitus: With ophthalmic complications | Diabetes Type 3 |
| E112 | Type 2 diabetes mellitus: With renal complications | Diabetes Type 4 |
| E119 | Type 2 diabetes mellitus: Without complications | Diabetes Type 5 |
| E143 | Unspecified diabetes mellitus: With ophthalmic complications | Diabetes unspecified |
| O249 | Diabetes mellitus in pregnancy, unspecified | Diabetes unspecified |
| R739 | Hyperglycaemia, unspecified | Diabetes, unspecified |
| E211 | Secondary hyperparathyroidism, not elsewhere classified | Endocrine |
| E201 | Pseudohypoparathyroidism | Endocrine |
| E210 | Primary hyperparathyroidism | Endocrine |
| E240 | Pituitary-dependent Cushing disease | Endocrine |
| E271 | Primary adrenocortical insufficiency | Endocrine |
| E213 | Hyperparathyroidism, unspecified | Endocrine, other |
| E221 | Hyperprolactinaemia | Endocrine, other |
| O244 | Diabetes mellitus arising in pregnancy | GDM |
| O100 | Pre-existing essential hypertension complicating pregnancy, childbirth and the puerperium | Hypertension |
| O11X | Pre-eclampsia superimposed on chronic hypertension | Hypertension |
| O141 | Severe pre-eclampsia | Hypertension |
| O149 | Pre-eclampsia, unspecified | Hypertension |
| O150 | Eclampsia in pregnancy | hypertension - eclampsia/pre-eclampsia |
| O152 | Eclampsia in the puerperium | hypertension - eclampsia/pre-eclampsia |
| O159 | Eclampsia, unspecified as to time period | hypertension - eclampsia/pre-eclampsia |
| O130 | Gestational [pregnancy-induced] hypertension | Hypertension - PIH |
| O13X | Gestational [pregnancy-induced] hypertension | Hypertension - PIH |
| I100 | Essential (primary) hypertension | hypertension (pre-existing) |
| I10X | Essential (primary) hypertension | hypertension (pre-existing) |
| O142 | HELLP syndrome | Hypertension, PET/eclampsia |
| O160 | Unspecified maternal hypertension | Hypertension, unspecified |
| O16X | Unspecified maternal hypertension | Hypertension, unspecified |
| A059 | Bacterial foodborne intoxication, unspecified | Infection |
| A084 | Viral intestinal infection, unspecified | Infection |
| A169 | Respiratory tuberculosis unspecified, without mention of bacteriological or histological confirmation | Infection |
| A401 | Sepsis due to streptococcus, group B | Infection |
| A415 | Sepsis due to other Gram-negative organisms | Infection |
| A419 | Sepsis, unspecified | Infection |
| A490 | Staphylococcal infection, unspecified site | Infection |
| B019 | Varicella without complication | Infection |
| B169 | Acute hepatitis B without delta-agent and without hepatic coma | Infection |
| B171 | Acute hepatitis C | Infection |
| B349 | Viral infection, unspecified | Infection |
| B54X | Unspecified malaria | Infection |
| B951 | Streptococcus, group B, as the cause of diseases classified to other chapters | Infection |
| B952 | Streptococcus group D and enterococcus as the cause of diseases classified to other chapters | Infection |
| B953 | Streptococcus pneumoniae as the cause of diseases classified to other chapters | Infection |
| B956 | Staphylococcus aureus as the cause of diseases classified to other chapters | Infection |
| J029 | Acute pharyngitis, unspecified | Infection |
| J039 | Acute tonsillitis, unspecified | Infection |
| J151 | Pneumonia due to Pseudomonas | Infection |
| J189 | Pneumonia, unspecified | Infection |
| K352 | Acute appendicitis with generalized peritonitis | Infection |
| K353 | Acute appendicitis with localized peritonitis | Infection |
| K358 | Acute appendicitis, other and unspecified | Infection |
| K358 | Acute appendicitis, other and unspecified | Infection |
| K37X | Unspecified appendicitis | Infection |
| K810 | Acute cholecystitis | Infection |
| K810 | Acute cholecystitis | Infection |
| K819 | Cholecystitis, unspecified | Infection |
| L030 | Cellulitis of finger and toe | Infection |
| L031 | Cellulitis of other parts of limb | Infection |
| L033 | Cellulitis of trunk | Infection |
| L038 | Cellulitis of other sites | Infection |
| L039 | Cellulitis, unspecified | Infection |
| L83X | Acanthosis nigricans | Infection |
| N136 | Pyonephrosis | Infection |
| N390 | Urinary tract infection, site not specified | Infection |
| N751 | Abscess of Bartholin gland | Infection |
| N764 | Abscess of vulva | Infection |
| O234 | Unspecified infection of urinary tract in pregnancy | Infection |
| O850 | Puerperal sepsis | Infection |
| O85X | Puerperal sepsis | Infection |
| O862 | Urinary tract infection following delivery | Infection |
| O984 | Viral hepatitis complicating pregnancy, childbirth and the puerperium | Infection |
| O989 | Unspecified maternal infectious or parasitic disease complicating pregnancy, childbirth and the puerperium | Infection |
| Q630 | Accessory kidney | Infection |
| Q831 | Accessory breast | Infection |
| R572 | Septic shock | Infection |
| R651 | Systemic Inflammatory Response Syndrome of infectious origin with organ failure | Infection |
| A600 | Herpes viral infection of genitalia and urogenital tract | Infection |
| A600D | Herpes viral infection of genitalia and urogenital tract | Infection |
| L732 | Hidradenitis suppurativa | infection |
| A539 | Syphilis, unspecified | Other medical |
| C770 | Secondary and unspecified malignant neoplasm: Lymph nodes of head, face and neck | Other medical |
| C774 | Secondary and unspecified malignant neoplasm: Inguinal and lower limb lymph nodes | Other medical |
| C786 | Secondary malignant neoplasm of retroperitoneum and peritoneum | Other medical |
| C851 | B-cell lymphoma, unspecified | Other medical |
| C910 | Acute lymphoblastic leukaemia [ALL] | Other medical |
| D561 | Beta thalassaemia | Other medical |
| D563 | Thalassaemia trait | Other medical |
| D569 | Thalassaemia, unspecified | Other medical |
| D571 | Sickle-cell anaemia without crisis | Other medical |
| E063 | Autoimmune thyroiditis | Other medical |
| E230 | Hypopituitarism | other medical |
| E272 | Addisonian crisis | Other medical |
| F501 | Atypical anorexia nervosa | Other medical |
| F502 | Bulimia nervosa | Other medical |
| F503 | Atypical bulimia nervosa | Other medical |
| G405 | Special epileptic syndromes | Other medical |
| G932 | Benign intracranial hypertension | Other medical |
| I620 | Subdural haemorrhage (acute)(nontraumatic) | Other medical |
| I64X | Stroke, not specified as haemorrhage or infarction | Other medical |
| I679 | Cerebrovascular disease, unspecified | Other medical |
| I712 | Thoracic aortic aneurysm, without mention of rupture | Other medical |
| I730 | Raynaud syndrome | Other medical |
| I776 | Arteritis, unspecified | Other medical |
| J47X | Bronchiectasis | Other medical |
| K510 | Ulcerative (chronic) pancolitis | Other medical |
| K519 | Ulcerative colitis, unspecified | Other medical |
| K754 | Autoimmune hepatitis | Other medical |
| K859 | Acute pancreatitis, unspecified | Other medical |
| L405 | Arthropathic psoriasis | Other medical |
| L405D | Arthropathic psoriasis | Other medical |
| L409 | Psoriasis, unspecified | Other medical |
| M029 | Reactive arthropathy, unspecified | Other medical |
| M059 | Seropositive rheumatoid arthritis, unspecified | Other medical |
| M060 | Seronegative rheumatoid arthritis | Other medical |
| M069 | Rheumatoid arthritis, unspecified | Other medical |
| M0690 | Rheumatoid arthritis, unspecified | Other medical |
| M074 | Arthropathy in Crohn disease [regional enteritis] | Other medical |
| M139 | Arthritis, unspecified | Other medical |
| M1390 | Arthritis, unspecified | Other medical |
| M1395 | Arthritis, unspecified | Other medical |
| M1396 | Arthritis, unspecified | Other medical |
| M1399 | Arthritis, unspecified | Other medical |
| M321 | Systemic lupus erythematosus with organ or system involvement | Other medical |
| M329 | Systemic lupus erythematosus, unspecified | Other medical |
| M349 | Systemic sclerosis, unspecified | Other medical |
| M350 | Sicca syndrome [Sjögren] | Other medical |
| M352 | Behçet disease | Other medical |
| M45X | Ankylosing spondylitis | Other medical |
| M45X0 | Ankylosing spondylitis | Other medical |
| M45X9 | Ankylosing spondylitis | Other medical |
| O222 | Superficial thrombophlebitis in pregnancy | Other medical |
| O870 | Superficial thrombophlebitis in the puerperium | Other medical |
| O981 | Syphilis complicating pregnancy, childbirth and the puerperium | Other medical |
| Q059 | Spina bifida, unspecified | Other medical |
| Q760 | Spina bifida occulta | Other medical |
| R760 | Raised antibody titre | Other medical |
| E039 | Hypothyroidism, unspecified | Thyroid disorder |
| E050 | Thyrotoxicosis with diffuse goitre | Thyroid disorder |
| E059 | Thyrotoxicosis, unspecified | Thyroid disorder |
| E069 | Thyroiditis, unspecified | Thyroid disorder |
